# Cortical hemodynamic dysregulation is linked to peripheral NIMETOX pathways in major depressive disorder: an 89-channel fNIRS systems-biology study

**DOI:** 10.64898/2026.09.28.26364187

**Authors:** Yiping Luo, Xia Deng, Mengqi Niu, Andre F Carvalho, Abbas F. Almulla, Hongzhou Wu, Xu Wang, Jing Li, Yingqian Zhang, Michael Maes

## Abstract

**Background:** Major depressive disorder (MDD) is clinically heterogeneous, but relationships among cortical hemodynamic abnormalities, peripheral neuroimmune, metabolic, and oxidative stress (NIMETOX) processes, and clinical phenome features remain unclear. We applied a systems-biology approach integrating functional near-infrared spectroscopy (fNIRS), peripheral NIMETOX biomarkers, and multidimensional clinical phenomes.

**Methods:** Patients with MDD (n = 86) and demographically matched healthy controls (HC, n = 40) underwent 89-channel high-density fNIRS during resting-state and verbal fluency task (VFT) conditions. Temporal hemodynamic variability (Difference) and area under the concentration–time curve (AUC) were derived from oxygenated hemoglobin (HbO), deoxygenated hemoglobin (HbR), and total hemoglobin (HbT) signals. NIMETOX assessments included inflammatory biomarkers, generalized and lipophilic antioxidant defenses, oxidized lipids, lipidomics, and 16S rRNA amplicon sequencing of the stool microbiome.

**Results:** MDD showed reduced VFT Difference and AUC across HbO, HbR, and HbT. Difference reductions were widespread across prefrontal, frontal eye field, language-related, and sensorimotor cortices, whereas AUC abnormalities were more restricted and resting-state alterations were limited. VFT differences significantly differentiated MDD from HC, with further improvement after integrating NIMETOX biomarkers. Lower antioxidants, carnitines/acetylcarnitines, polyunsaturated and even-chain saturated fatty acids, inflammation, indices of gut dysbiosis and leaky gut, and adverse childhood experiences explained substantial variance in HbO, HbR, and HbT Difference. Integrated fNIRS– NIMETOX profiles explained substantial variance in overall illness severity (cross-validated R² = 0.62) and physiosomatic symptoms (cross-validated R² = 0.48).

**Conclusions:** Systems-biology integration identified a multilevel MDD phenotype linking peripheral NIMETOX dysregulation with task-evoked cortical hemodynamic abnormalities and clinical phenome heterogeneity, supporting an interconnected gut-NIMETOX-brain pathophysiology.

**Highlights:**

- MDD is characterized by reduced temporal hemodynamic variability and impaired task-evoked recruitment across oxygenated (HbO) and deoxygenated (HbR) hemoglobin.
- Multidimensional fNIRS features discriminate MDD from controls, with further improvement after integration with peripheral neuroimmune, metabolic and oxidative stress (NIMETOX) pathways.
- Peripheral NIMETOX biomarkers, particularly lipophilic antioxidants, oxidized lipids, inflammation, and gut-microbiome related indices, explain substantial variation in HbO/HbR hemodynamic abnormalities and the clinical features of MDD.
- Multilevel systems-biological aberrations link peripheral NIMETOX pathways with brain hemodynamic abnormalities and the symptoms of MDD.

## 1. Introduction

Contemporary psychiatric classifications remain largely symptom-based, with limited incorporation of objective biological mechanisms, thereby constraining their biological validity^[1,2]^. Moreover, clinical, molecular, and neuroimaging findings are typically analyzed separately, rather than as components of an integrated pathophysiological system^[3]^. Nomothetic network psychiatry addresses this fragmentation by integrating clinical phenomes, peripheral biomarkers, and neuroimaging features within a bottom-up, data-driven systems-biology framework^[3,4]^. Such multilevel integration may identify biological pathways underlying dimensional psychopathology and link systemic dysregulation to brain dysfunction and the clinical expression of major depressive disorder (MDD)^[3]^.

Functional near-infrared spectroscopy (fNIRS) enables non-invasive, continuous assessment of cortical hemodynamics by estimating relative changes in oxygenated hemoglobin (HbO), deoxygenated hemoglobin (HbR), and total hemoglobin (HbT)^[5,6]^. These hemodynamic signals reflect complementary aspects of oxygenated blood supply, oxygen extraction and metabolism, and cerebral blood volume^[7]^. Different response patterns may indicate distinct disturbances; for example, reduced HbO with increased HbT may reflect inefficient microvascular recruitment, whereas reduced HbO with relatively stable HbR may indicate insufficient perfusion or vasomotor recruitment^[8,9]^. Previous fNIRS studies have consistently reported attenuated task-evoked prefrontal and frontotemporal HbO responses in MDD, particularly during verbal fluency tasks (VFTs)^[5,10]^. However, most studies have focused on mean HbO or single activation-amplitude measures, with fewer simultaneously examining HbO, HbR, and HbT across resting and task conditions^[5,11]^. Conventional mean concentrations and general linear model (GLM) estimates may also overlook temporal information contained within the hemodynamic response^[12,13]^. Area under the concentration–time curve (AUC) quantifies cumulative hemodynamic output, whereas “Difference,” defined as the mean absolute change between successive time points, captures short-term temporal hemodynamic variability^[13,14]^. Their combined assessment may therefore provide a more comprehensive characterization of cortical hemodynamic dysfunction^[15]^. fNIRS abnormalities in MDD have been associated with symptom severity, cognitive performance, treatment response, and clinical state^[5,16,17]^. Thus, multidimensional fNIRS may characterize both regional cortical dysfunction and clinically relevant alterations in neurovascular dynamics in MDD.

Importantly, MDD-related cortical hemodynamic abnormalities should not be considered in isolation from systemic pathophysiology. Increasing evidence indicates that MDD involves interactions between peripheral neuroimmune, metabolic, and oxidative stress (NIMETOX) pathways and the central nervous system (CNS)^[18,19]^. A key peripheral biological feature of MDD is activation of the acute-phase inflammatory (API) response, characterized by increased serum monomeric C-reactive protein (mCRP) together with reduced levels of the negative acute-phase proteins albumin and transferrin^[20]^. This inflammatory response is accompanied by impaired antioxidant defenses, including total antioxidant capacity (TAOC), apolipoprotein A1 (ApoA1), paraoxonase-1 (PON1) activity, and high-density lipoprotein cholesterol (HDL-C)[21]. More specifically, our recent work demonstrated increased oxidized high-density lipoprotein (OxHDL), together with reduced HDL-C and oxidized low-density lipoprotein (OxLDL), resulting in a shift toward preferential oxidative modification of HDL particles. These findings suggest that high-density lipoprotein (HDL) particles may be particularly vulnerable to oxidative and inflammatory injury in MDD, with consequent impairment of HDL-associated antioxidant and endothelial-protective functions^[21]^.

These NIMETOX pathways may be driven partly by gut dysbiosis with increased intestinal permeability (leaky gut)^[18,22]^, facilitating lipopolysaccharide (LPS) translocation and consequent immune-inflammatory and oxidative responses via for example the Toll-like receptor (TLR)-radical cycle^[23–25]^. Gut dysbiosis may additionally alter short-chain fatty-acid (SCFA) metabolism, with lower levels of protective SCFAs, including acetate, propionate, and butyrate^[26]^. Moreover, this gut–NIMETOX dysregulation is accompanied by extensive lipid and fatty-acid remodeling, including reduced even-chain saturated fatty acids (ECSFAs), polyunsaturated fatty acids (PUFAs), very-long-chain fatty acids (VLCFAs), carnitine/sphingomyelin module (CARSM) and ether lipids, together with an increased ceramide-, GM3 ganglioside-, N-acyl-phosphatidylethanolamine-, and phosphatidylethanolamine-enriched lipid component (CERLNAPE)^[23,27]^. Impaired reverse cholesterol transport (RCT), reflected by a reduced composite RCT index incorporating lecithin–cholesterol acyltransferase (LCAT) activity, HDL-C, and ApoA1, constitutes an additional component of lipid dysregulation in MDD. Reduced RCT capacity may interact with the acute-phase inflammatory response and impaired lipophilic antioxidant defenses, thereby linking disturbances in cholesterol handling with the broader neuroimmune–metabolic– oxidative stress pathology of MDD^[20,28]^.

Collectively, these alterations define a lipid-depletion (PUFAs, ECSFAs, VLCFAs, ether lipids, carnitines/acetylcarnitines, ApoA1, HDL-C) – lipotoxicity (increased ceramides and GM3 gangliosides) phenotype of MDD, which is further shaped by acute-phase inflammatory responses, lowered antioxidant defenses and RCT, and increased oxidative stress. These peripheral abnormalities may affect neurovascular-unit homeostasis through endothelial function, cerebral blood-flow regulation, and mitochondrial energy metabolism, thereby providing a potential mechanistic explanation for cortical hemodynamic abnormalities in MDD^[29,30]^. Therefore, cortical hemodynamic changes measured by fNIRS may provide a quantifiable window through which to examine associations between peripheral biological abnormalities and CNS functions^[7,18,31]^.

However, MDD studies have largely examined clinical symptoms, peripheral biology, and brain function separately, with few integrating multidimensional phenomes, NIMETOX features, and fNIRS hemodynamics^[5,32,33]^. Consequently, it remains unclear whether peripheral NIMETOX processes explain individual differences in cortical hemodynamics and whether both jointly explain key clinical phenomes, including illness severity, physiosomatic symptoms, and recurrence of illness (RECUR)^[5,34,35]^. Furthermore, few studies have evaluated the discriminative value of these multidimensional hemodynamic features for distinguishing MDD from healthy individuals or examined whether their integration with peripheral NIMETOX features improves the explanation of current symptom severity and subclinical phenotypic variation**^[36,37]^.**

Hence, the present study aimed to: (a) characterize a multidimensional cortical hemodynamic phenotype in MDD inpatients using 89-channel fNIRS, integrating resting-state and VFT conditions, HbO/HbR/HbT signals, and two complementary time-domain measures, Difference and AUC; and (b) integrate this CNS phenotype with peripheral gut-NIMETOX pathways and key clinical phenomes. First, we compared hemodynamic features between MDD and healthy controls (HC) and characterized their spatial distribution using region of interest (ROI) analyses. Second, we examined associations between peripheral gut-NIMETOX features and fNIRS phenotypes. Third, we investigated how fNIRS hemodynamic components, coupled with gut-NIMETOX features, explain key clinical phenomes, including overall severity of depression (OSOD), cognitive-affective and physiosomatic symptoms, and RECUR.

## Materials and Methods

### Participants and study design

A case-control cross-sectional design was adopted. A total of 126 right-handed participants were recruited from the Psychosomatic Medicine Center of Sichuan Provincial People’s Hospital, Chengdu, China, including 40 HC and 86 inpatients with MDD. All inpatients were diagnosed with MDD by a senior psychiatrist through structured clinical interviews according to the Diagnostic and Statistical Manual of Mental Disorders, Fifth Edition (DSM-5)^[38]^. Additional inclusion criteria were as follows: (a) age 18–65 years; (b) right-handedness confirmed using the Edinburgh Handedness Inventory; (c) completion of at least six years of formal education to ensure adequate comprehension of cognitive assessments; and (d) no clinically significant structural brain abnormalities on magnetic resonance imaging (MRI). Healthy controls comprised community volunteers and hospital staff and were group-matched to the MDD group for age, sex, years of education, and body mass index (BMI). They had no lifetime history of DSM-5 psychiatric disorders and no family history of MDD, bipolar disorder, substance use disorders, or suicide. We excluded participants with the following conditions: (a) other major psychiatric or neurodevelopmental disorders, including schizophrenia, bipolar disorder, schizoaffective disorder, eating disorders, autism spectrum disorder, or neurocognitive disorders; (b) neurological disorders or a history of brain injury, including epilepsy, stroke, multiple sclerosis, Alzheimer’s disease, Parkinson’s disease, brain tumors, or traumatic brain injury; (c) severe cardiovascular, hepatic, renal, or endocrine diseases; (d) severe systemic, autoimmune, inflammatory, or malignant diseases, or a recent severe infection; (e) pregnancy or lactation; (f) a history of non-medical substance or alcohol abuse or dependence, excluding nicotine dependence; and (g) severe cognitive impairment, contraindications to fNIRS, or inability to complete the experimental task.

All participants or their legal representatives provided written informed consent after a thorough explanation of the study’s objectives, procedures, and potential risks. The study protocol was formally approved by the Ethics Committee of Sichuan Provincial People’s Hospital [Ethics (Research) 2024–203]. The a priori sample size was estimated before data analysis using G*Power version 3.1.9.4^[39]^ for a fixed-model linear multiple regression (the primary statistical analysis with OSOD as outcome variable) test of R² deviation from zero. Assuming that the model would explain 15.0% of the outcome variance (R² = 0.15; f² = 0.176), with an alpha level of 0.05, statistical power of 0.80, and a maximum of seven predictors, the minimum required total sample size was 89 participants. The final analytical cohort comprised 124 participants, and the smallest model-specific complete-case sample comprised 102 participants; both exceeded the calculated minimum requirement, supporting the adequacy of the multivariable regression analyses.

### Clinical assessment and phenotyping

Clinical phenotyping was conducted for all participants. A senior psychiatrist collected demographic information, medical history, psychosocial and developmental history, and family psychiatric history through structured interviews. Current and lifetime DSM-5 psychiatric disorders were assessed using the Mini-International Neuropsychiatric Interview (M.I.N.I.)^[40]^. On the day of fNIRS acquisition, clinician-rated depressive and anxiety symptom severity was assessed using the 21-item Hamilton Depression Rating Scale (HAMD-21) and the Hamilton Anxiety Rating Scale (HAMA), respectively^[41,42]^. Self-reported depressive symptoms and somatic symptom burden were evaluated using the Beck Depression Inventory (BDI) and the Somatic Symptom Scale-8 (SSS-8), respectively^[43–45]^. State anxiety and fatigue-related physiosomatic symptoms were additionally assessed using the State subscale of the State-Trait Anxiety Inventory (STAI-State) and the 12-item FibroFatigue Scale (FFS)^[46,47]^, respectively. Lifetime and current suicidal ideation and suicide attempts were assessed using the Columbia-Suicide Severity Rating Scale (C-SSRS)^[48]^. These clinical measures were used to construct the key phenome indices, including OSOD, the physiosomatic phenome, and RECUR. The RECUR composite integrated the standardized number of depressive episodes, lifetime suicidal ideation, and lifetime suicide attempts, as explained previously^[23,25–27]^. Pure BDI was computed by summing only the cognitive-affective BDI symptoms, excluding all somatic symptoms. All clinical assessments were administered or supervised by the same senior psychiatrist on the day of fNIRS acquisition to ensure procedural consistency.

Anthropometric and metabolic assessments were performed as previously described^[49]^. Body mass index (BMI) was calculated as weight (kg) divided by height squared (m²), and waist circumference (WC) was measured midway between the lowest rib and the iliac crest. Metabolic syndrome (MetS) was defined according to the 2009 Joint Scientific Statement criteria, requiring the presence of at least three of the following five components: elevated WC (≥90 cm in men or ≥80 cm in women), triglycerides ≥150 mg/dL, reduced HDL cholesterol (<40 mg/dL in men or <50 mg/dL in women), blood pressure ≥130/85 mmHg or antihypertensive treatment, and fasting glucose ≥100 mg/dL or a diagnosis of diabetes^[50]^.

### NIMETOX assays

Electronic Supplementary File (ESF), Table 1 lists all NIMETOX biomarkers measured in the current study and the methods used to quantify them. ESF, Table 2 lists the different NIMETOX biomarkers and composites used in the current study.

### fNIRS data acquisition

fNIRS was used to quantify cortical hemodynamic responses during resting-state and VFT conditions^[6,10]^. To minimize environmental and motion-related artifacts, we conducted data acquisition in a sound-attenuated room. Participants were instructed to remain relaxed and minimize head and facial movements throughout the recording. Signals were acquired using an 89-channel continuous-wave fNIRS system (BS-2000, Wuhan Zilian Hongkang Co., Ltd., China). The optode array provided broad coverage of the frontal, parietal, temporal, and temporoparietal cortices and comprised 27 dual-wavelength near-infrared light sources operating at 690 and 830 nm and 25 detectors arranged in a multichannel grid.

The optode array was positioned symmetrically on the scalp according to the international 10–20 system, with source optode S2 aligned with Fpz. A three-dimensional electromagnetic digitizer (NirMap, Wuhan Zilian Hongkang Co., Ltd., Wuhan, China) was used to record the spatial coordinates of the principal cranial landmarks (Nz, Cz, AL, and RL) and all individual optodes. Channel coordinates were probabilistically registered to the Montreal Neurological Institute (MNI) standard brain using the near-infrared spectroscopy statistical parametric mapping (NIRS-SPM) spatial registration framework and mapped to the corresponding cortical areas^[51,52]^.

Based on this spatial registration, 84 of the 89 channels were assigned to 16 lateralized ROIs, representing eight bilateral cortical regions: the dorsolateral prefrontal cortex (DLPFC), frontopolar area (FPA), frontal eye fields (FEF), temporal cortex (TC), premotor and supplementary motor cortex (PreM/SMC), primary motor cortex (M1), Broca’s area and its right-hemispheric homolog, and primary somatosensory cortex (S1). The five midline channels were not included in the lateralized ROI aggregation. Detailed channel-to-ROI assignments are provided in ESF, Table 3.

### Data preprocessing pipeline

fNIRS data were processed using NirMaster data analysis software, version V1.1 (Wuhan Yiruide Medical Equipment New Technology Co., Ltd., Wuhan, China), according to the following sequential pipeline: (1) Downsampling: The raw signals were downsampled to 10 Hz to standardize the sampling rate and reduce high-frequency signal components. (2) Channel- and participant-level quality control: The coefficient of variation (CV), calculated as the standard deviation divided by the mean, was determined for each channel. Channels with a CV > 15% were classified as bad channels. Participants with at least 27 bad channels, corresponding to ≥30% of the 89-channel montage, were excluded from the primary fNIRS analytical cohort. (3) Artifact correction and filtering: Raw optical intensity signals were first converted to optical density (OD). Motion artifacts were identified using a 0.5-s detection window (TMOTION), a standard-deviation threshold of 20.0 (STDE), an amplitude threshold of 0.50 (AMP), and a 1.0-s masking window (TMASK), and were subsequently corrected using spline interpolation. A band-pass filter of 0.01–0.1 Hz was then applied to attenuate cardiac, respiratory, and very-low-frequency baseline components. (4) Hemoglobin concentration conversion: Changes in OD were converted into dynamic concentration-change time series for HbO, HbR, and HbT according to the modified Beer–Lambert law (MBLL)^[6]^. (5) VFT epoching, block averaging, and baseline correction: For task-related analyses, epochs encompassing the 10-s pre-task baseline, active task period, and post-task recovery period were extracted. A linear baseline trend was estimated using the 10-s pre-task and 5-s post-task segments to correct for baseline drift. The mean value of the 10-s pre-task baseline was subsequently subtracted, after which the task epochs were block-averaged. Finally, a 5-s moving-average window was applied to smooth the signals. Resting-state data were retained as continuous preprocessed time series for subsequent segmentation and feature extraction. (6) For retained participants, missing ROI-level values were imputed using a predefined neighbor-averaging procedure based on nearby valid observations. (7) Segmentation and feature extraction: The preprocessed recordings were segmented into resting-state and VFT periods according to the experimental protocol. For the HbO, HbR, and HbT time series, the AUC and the Difference metric were extracted separately from each segment. Difference was defined as the mean of the absolute differences between each pair of consecutive data points within the specified time window and was used as an index of short-term, point-to-point hemodynamic variability. Resting-state and task-derived AUC and Difference features were entered into subsequent statistical analyses.

### Statistical analysis

#### Analysis of demographic and clinical variables

IBM SPSS Statistics for Windows, Version 30.0 (IBM Corp., Armonk, NY, USA), was used for all clinical and demographic statistical analyses. All tests were two-sided, with the significance level set at α = 0.05. Sex and dichotomized marital status were compared using Pearson’s chi-square test. Analysis of variance and GLM analysis were used to compare scale variables between diagnostic categories. **Figure 1** provides an overview of the machine-learning approaches employed in the data analysis.

**Figure 1.**
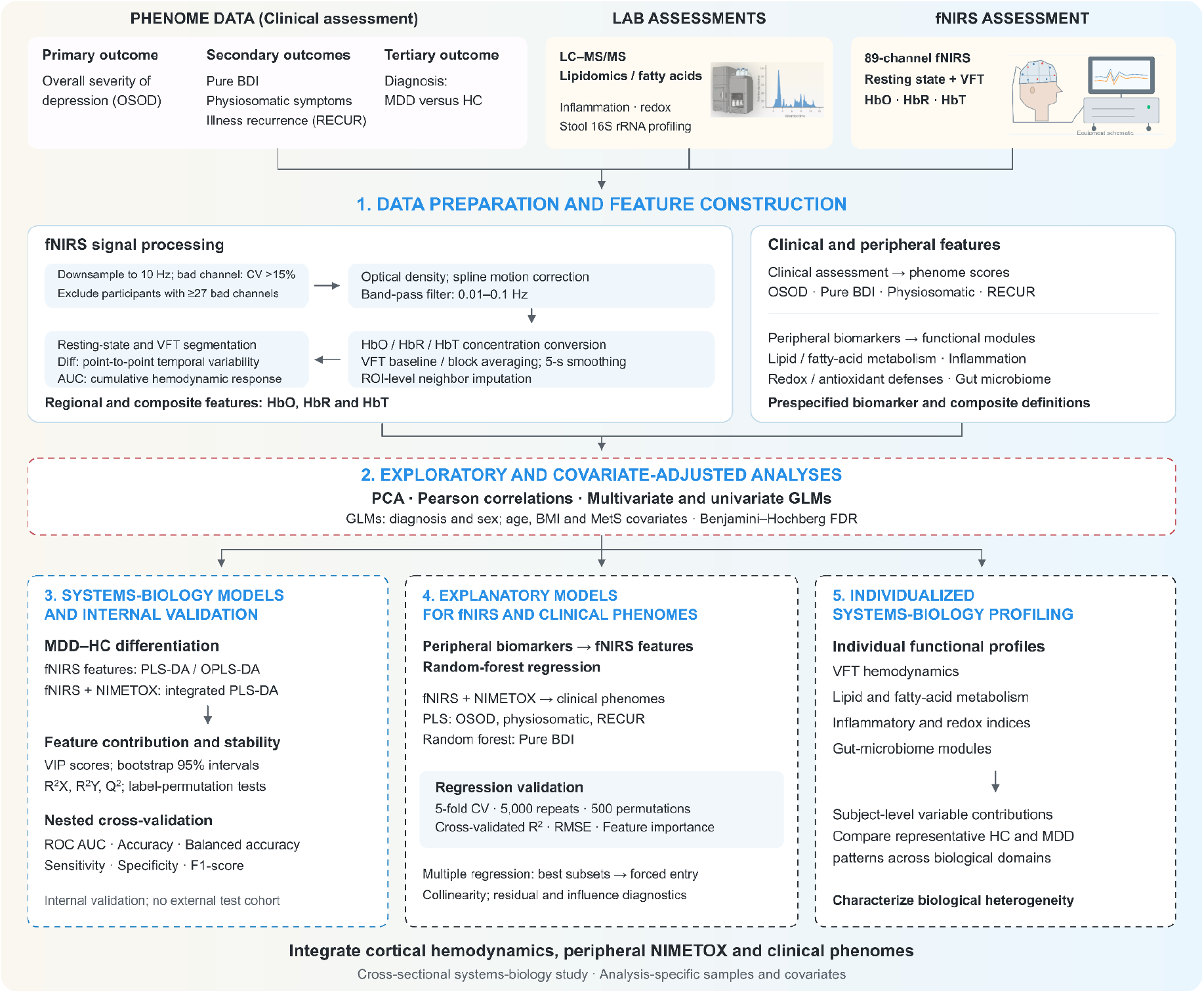
Summary of the machine-learning and systems-biology workflow used to integrate clinical, fNIRS, and peripheral NIMETOX data. Following quality control and preprocessing, resting-state and VFT HbO, HbR, and HbT features were extracted. PCA and correlation analyses were used for exploratory characterization of fNIRS features and their relationships with selected NIMETOX biomarker modules. OPLS-DA and PLS-DA were applied for multivariate differentiation of patients with major depressive disorder (MDD) and healthy controls and integrated feature selection. Model robustness was assessed using permutation testing and nested cross-validation. Individualized NIMETOX profiles were subsequently constructed to characterize biological heterogeneity within MDD. Multiple regression examined adjusted associations between fNIRS components, clinical variables, and peripheral biological features. Random-forest regression quantified the cross-validated variance in HbO, HbR, and HbT VFT Difference explained by peripheral NIMETOX features. PLS regression integrated fNIRS and NIMETOX features to explain variation in the key phenome, namely OSOD. Together, these analyses provided an internally validated framework linking peripheral NIMETOX dysregulation, cortical hemodynamic abnormalities, and the MDD clinical phenome.

#### Exploratory principal component analysis

ROI-level HbO, HbR, and HbT features during the VFT were examined using unsupervised principal component analysis (PCA) to assess the overall distribution of MDD and HC in a low-dimensional feature space.

#### Hemodynamic differentiation of MDD and HC

To examine cerebral hemodynamic differences between MDD and HC, covariate-adjusted multivariate and univariate GLMs were fitted separately for the VFT and resting-state conditions. Diagnostic group and sex were entered as fixed factors, with age, MetS, and BMI as covariates. Six standardized fNIRS indices were examined: Difference and AUC for HbO, HbR, and HbT. Benjamini– Hochberg false discovery rate (FDR) correction was applied to control for multiple comparisons, with q < 0.05 considered significant^[53]^.

Supervised MDD–HC differentiation based on fNIRS features was examined using partial least-squares discriminant analysis (PLS-DA) and orthogonal partial least-squares discriminant analysis (OPLS-DA)^[54]^. Model fit and predictive performance were assessed using R²X, R²Y, and Q², while label-permutation testing was used to evaluate model stability and potential overfitting. Variable importance in projection (VIP) scores were used to identify the HbO, HbR, and HbT Difference features contributing most strongly to group differentiation. To determine whether peripheral biological information provided additional discriminative information, VFT Difference features were subsequently integrated with peripheral biomarkers using PLS-DA. The model included candidate fNIRS and peripheral NIMETOX variables and was evaluated using nested cross-validation and label-permutation testing. Repeated bootstrap resampling was used to assess the stability of VIP contributions and derive 95% confidence intervals. Internal discriminative performance was assessed using receiver operating characteristic AUC, accuracy, sensitivity, specificity, balanced accuracy, and F1-score.

#### Explanation of fNIRS and clinical phenotypes

Random-forest (RF) regression^[55]^ was used to determine how well peripheral biomarkers explained the three Difference and AUC phenotypes (HbO, HbR, and HbT) during the VFT and resting state. Candidate predictors included metabolic, inflammatory, oxidative-stress, lipid and fatty-acid, SCFA, and gut-microbiota biomarkers. Predictive performance was assessed using 5-fold cross-validation repeated 5,000 times, while 500 permutation tests determined whether model performance exceeded that expected by chance. Performance was expressed as cross-validated R² and root mean square error (RMSE). Variable-importance analyses identified the peripheral biomarkers contributing most strongly to each model and those consistently important across the three hemoglobin measures. Partial least-squares (PLS) regression models were also constructed for OSOD, the physiosomatic phenome, and RECUR, whereas RF regression was used for Pure BDI, with ROI-level HbO, HbR, and HbT Difference features and NIMETOX biomarkers as explanatory variables.

Multiple linear regression analyses were used to examine associations between precomputed fNIRS hemodynamic component scores as dependent variables and the NIMETOX biomarkers and relevant covariates entered as explanatory variables. Toward this end we computed z unit-based composite scores based on the sums of all VFT (difference and AUC) HbO, HbR or HbT values. Model building was performed in SPSS using best-subsets regression combined with an overfitting-control algorithm to identify parsimonious predictor combinations with adequate model stability. Among the resulting candidate models, final subsets were selected considering statistical performance, parsimony, collinearity, and consistency with established systems-biology knowledge. The selected predictors were subsequently entered simultaneously into a conventional forced-entry multiple regression model to estimate their independent contributions and obtain the final regression coefficients and model statistics. For each final model, we report the model F statistic with degrees of freedom and p value and R². For each explanatory variable, standardized regression coefficients (β), t values, and exact p values were reported. Collinearity was evaluated using tolerance and variance inflation factor (VIF) statistics. Model assumptions were evaluated by inspecting residual-versus-predicted plots for heteroscedasticity and normal probability– probability (P–P) plots of standardized residuals for deviations from normality. Potential influential observations and outliers were examined using standardized residuals, leverage values, and Cook’s distance.

## Results

### 3.1 Demographic, clinical and NIMETOX characteristics

**Table 1** summarizes the demographic and clinical characteristics of the analytical sample. The two groups did not differ significantly in sex distribution, age, years of education, BMI, or MetS. In contrast, the MDD group had a substantially greater symptom burden than HC, with significantly higher OSOD, RECUR, Pure BDI, and physiosomatic phenome scores.

### 3.2 Exploratory analysis

**Figure 2A** shows the PCA results with the distribution of MDD and HC based on VFT Difference features. The PCA score plot showed a tendency toward group-related distribution, with substantial overlap between participants with MDD and HC. Principal component 1 (PC1) accounted for 57.6% of the total variance and showed the main tendency toward group separation, whereas principal component 2 (PC2) accounted for an additional 6.7%. MDD subjects were predominantly distributed toward positive PC1 scores, whereas HC showed a broader distribution toward negative PC1 scores. Nevertheless, some overlap between the groups remained, indicating that the multivariate profile differed between MDD and HC without complete separation.

**Figure 2.**
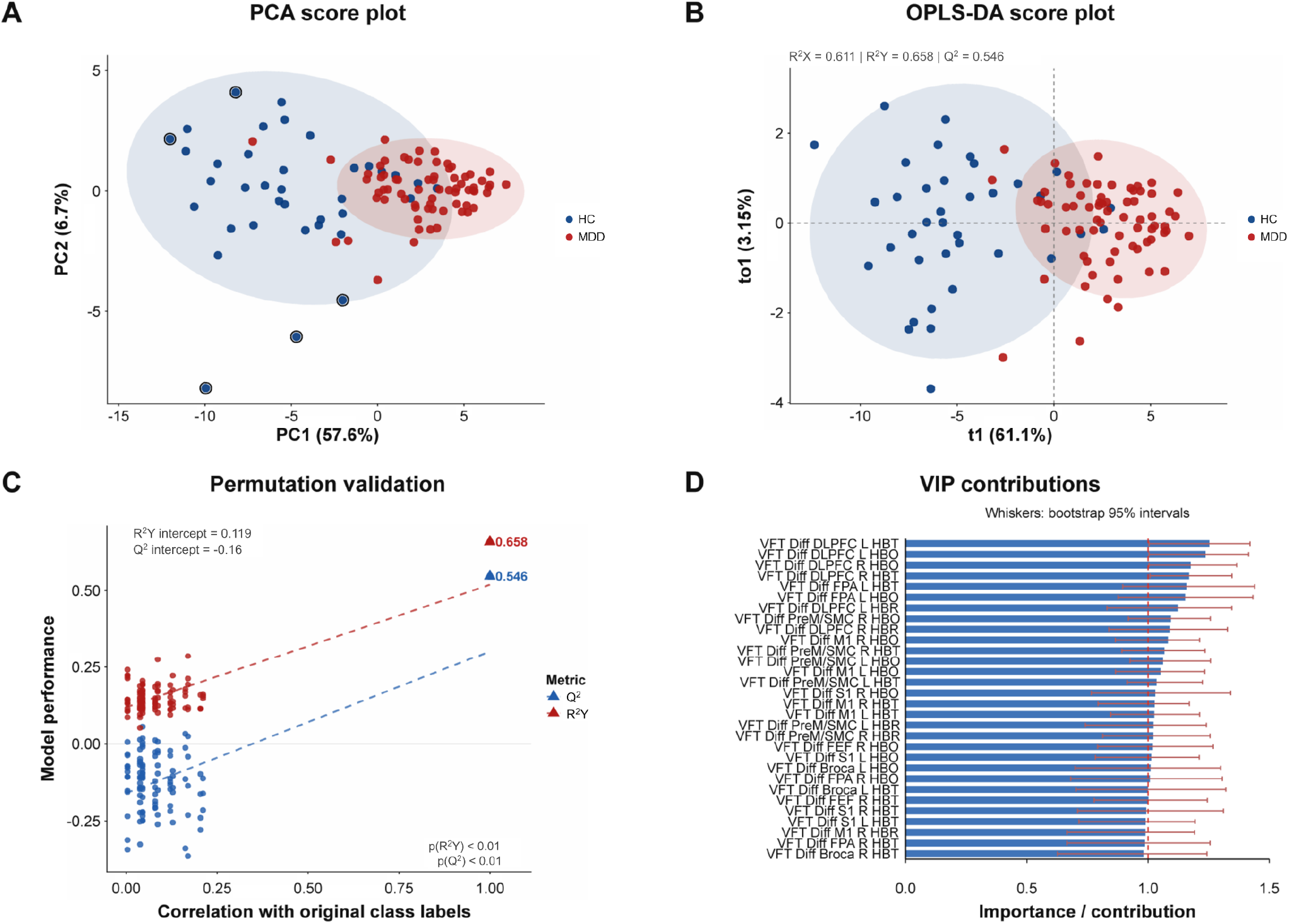
Multivariate differentiation of major depressive disorder (MDD) and healthy controls (HC) based on fNIRS VFT Difference features. (A) Principal component analysis (PCA) score plot showing the distribution of MDD and HC based on VFT Difference features. (B) OPLS-DA score plot showing supervised multivariate differentiation between MDD and HC. (C) Permutation testing of the OPLS-DA model. (D) Variable importance in projection (VIP) scores identifying the fNIRS features contributing most strongly to group differentiation. VFT, verbal fluency task; OPLS-DA, orthogonal partial least-squares discriminant analysis; VIP, variable importance in projection.

### 3.3 Univariate hemodynamic and NIMETOX alterations in MDD

Covariate-adjusted GLMs were used to compare fNIRS indices between the MDD and HC groups during resting-state and VFT conditions (**Table 2**). During resting state, the MDD group showed lower HbO, HbR, and HbT Difference values than the HC group; however, after FDR correction, only the reduction in HbO Difference remained significant. No significant group differences were observed in HbO, HbR, or HbT AUC. During the VFT, the MDD group showed significantly lower Difference and AUC values for HbO, HbR, and HbT than the HC group after FDR correction.

ESF, Table 4 shows that, after FDR correction, VFT Difference values for HbO, HbR, and HbT were significantly lower in the MDD group across the bilateral DLPFC, FPA, FEF, PreM/SMC, M1, S1, and Broca’s area. For VFT AUC, significantly lower HbO values were observed in the right PreM/SMC and left Broca’s area. Significantly lower HbR values were observed in the left PreM/SMC, bilateral M1, and right Broca’s area, whereas significantly lower HbT values were observed in the right PreM/SMC, left S1, and left Broca’s area. During resting state, no ROI-level group differences in Difference or AUC for HbO, HbR, or HbT remained significant after FDR correction.

Covariate-effect analyses showed significant effects of age on the VFT HbO Difference (p = 0.001), VFT HbR Difference (p = 0.002), and VFT HbT Difference (p = 0.002) models, as well as in all resting-state Difference and AUC models (all p < 0.001). Sex showed significant effects in the VFT HbR Difference (p < 0.001), resting-state HbO AUC (p = 0.009), and resting-state HbT AUC (p = 0.002) models. Neither MetS nor BMI showed significant effects in any model, even before FDR correction. In addition, there were no significant effects of the drug state variables on the fNIRS results (results of multivariate and univariate GLM analysis with age, sex and BMI as covariates (antidepressants: n = 62; benzodiazepines: n = 43; atypical antipsychotics: n = 31; mood stabilizers: n = 8)).

ESF, Table 5 shows the differences in the NIMETOX biomarkers among patients with MDD and controls. As described previously, no significant effects of the drug state were detected on any of these NIMETOX biomarkers[21,23,24,26,27].

### Multivariate systems-biology characterization using PLS-DA

OPLS-DA demonstrated multivariate differentiation between MDD and HC, with partial overlap between the groups (**Figure 2B–D**). The model yielded R²X = 0.611, R²Y = 0.658, and Q² = 0.546, indicating substantial explained variance and internally cross-validated model performance. Permutation testing supported model validity, with substantially lower R²Y and Q² values for permuted models and intercepts of 0.119 and −0.160, respectively (both permutation p < 0.01). VIP analysis identified predominantly DLPFC, FPA, and PreM/SMC hemodynamic measures among the major contributors to the multivariate group differentiation.

**Figure 3** shows the integrated systems-biology results obtained by combining peripheral biomarkers with the VFT Difference features. The integrated PLS-DA model yielded a nested cross-validated AUC of 0.982 and an accuracy of 0.958 (**Figure 3E**). Bootstrap analysis supported the stability of the VIP contributions. The variables showing the highest VIP contributions primarily comprised ECSFAs, VLCFAs, PUFA remodeling, DLPFC-related VFT Difference features, and the LPS-TLR module. At a distance, other ROIs, All antioxidants, and CARSM also showed VIP scores > 1. Compared with the model using Difference features alone, the addition of peripheral biomarkers further improved internal cross-validation performance, indicating that peripheral biological features contributed additional multivariate information beyond the fNIRS hemodynamic phenotype. These findings reflect internal model validity and require confirmation in an independent external cohort.

**Figure 3.**
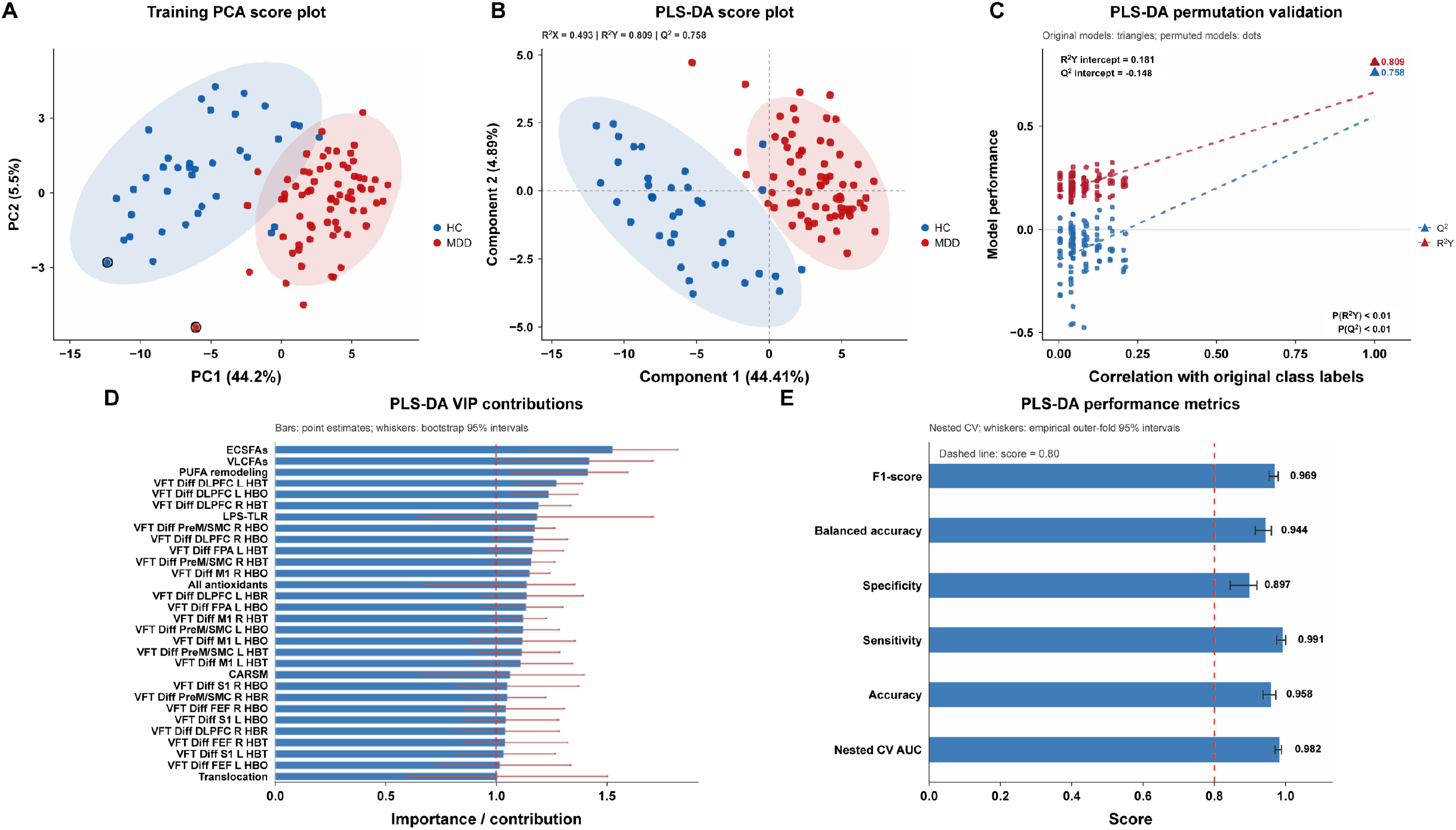
Integrated PLS-DA of fNIRS VFT Difference features and peripheral NIMETOX biomarkers. (A) PCA score plot of the integrated feature set. (B) PLS-DA score plot showing multivariate differentiation between MDD and HC. (C) Permutation validation of the PLS-DA model. (D) VIP contributions of fNIRS and peripheral NIMETOX variables; error bars indicate bootstrap 95% confidence intervals. (E) Nested cross-validation performance, including AUC, accuracy, sensitivity, specificity, balanced accuracy, and F1-score. PLS-DA, partial least-squares discriminant analysis; AUC, area under the receiver operating characteristic curve.

### 3.4 Associations of fNIRS hemodynamic phenotypes with peripheral biomarkers

**Figure 4** shows that HbO, HbR and HbT Difference composites were significantly correlated with NIMETOX biomarkers except the arachidonic acid/eicosapentaenoic acid (AA/EPA) ratio, butanoic acid, and Protective SCFAs. **Figure 5** shows that the RF regression models yielded cross-validated R² values of 0.44, 0.39, and 0.43 for HbO, HbR, and HbT Difference, respectively. The most consistently important predictors across models were ECSFAs, PUFA remodeling, CERLNAPE and VLCFAs, followed at a distance by CARSM, the API index, All antioxidants, and gut modules. These results show that peripheral NIMETOX biomarkers explained a substantial proportion of individual differences in hemodynamic dynamics during the VFT. These models indicate that peripheral NIMETOX biomarkers were associated with individual differences in the VFT Difference measures.

**Figure 4.**
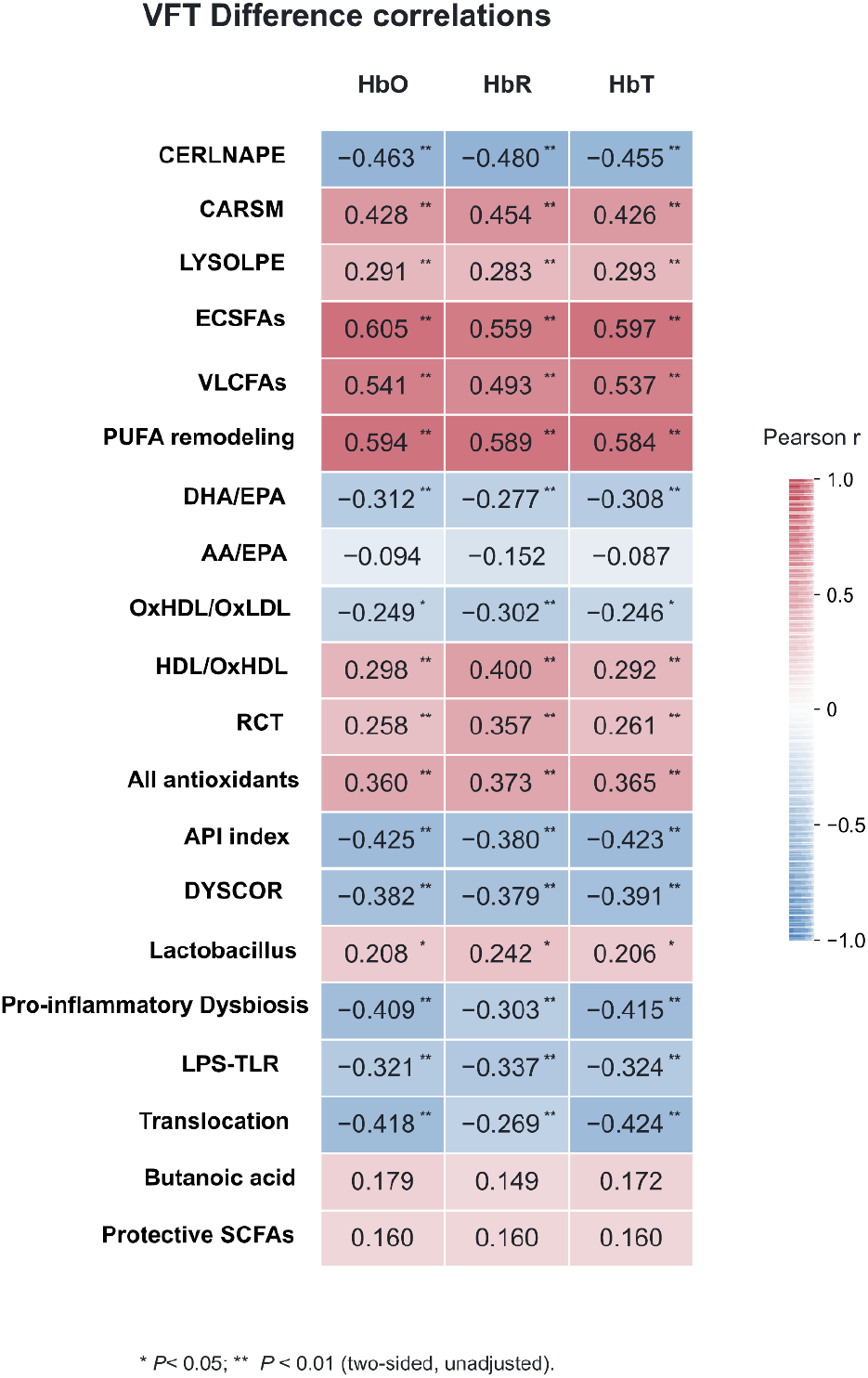
Associations between fNIRS VFT Difference phenotypes and peripheral NIMETOX biomarkers. Correlation matrix showing associations of HbO, HbR, and HbT VFT Difference composites with peripheral metabolic, inflammatory, oxidative-stress, lipid/fatty-acid, short-chain-fatty-acid, and gut-microbiome variables. HbO, oxygenated hemoglobin; HbR, deoxygenated hemoglobin; HbT, total hemoglobin. See ESF, Tables 1 and 2 for a detailed description of the NIMETOX biomarkers.

**Figure 5.**
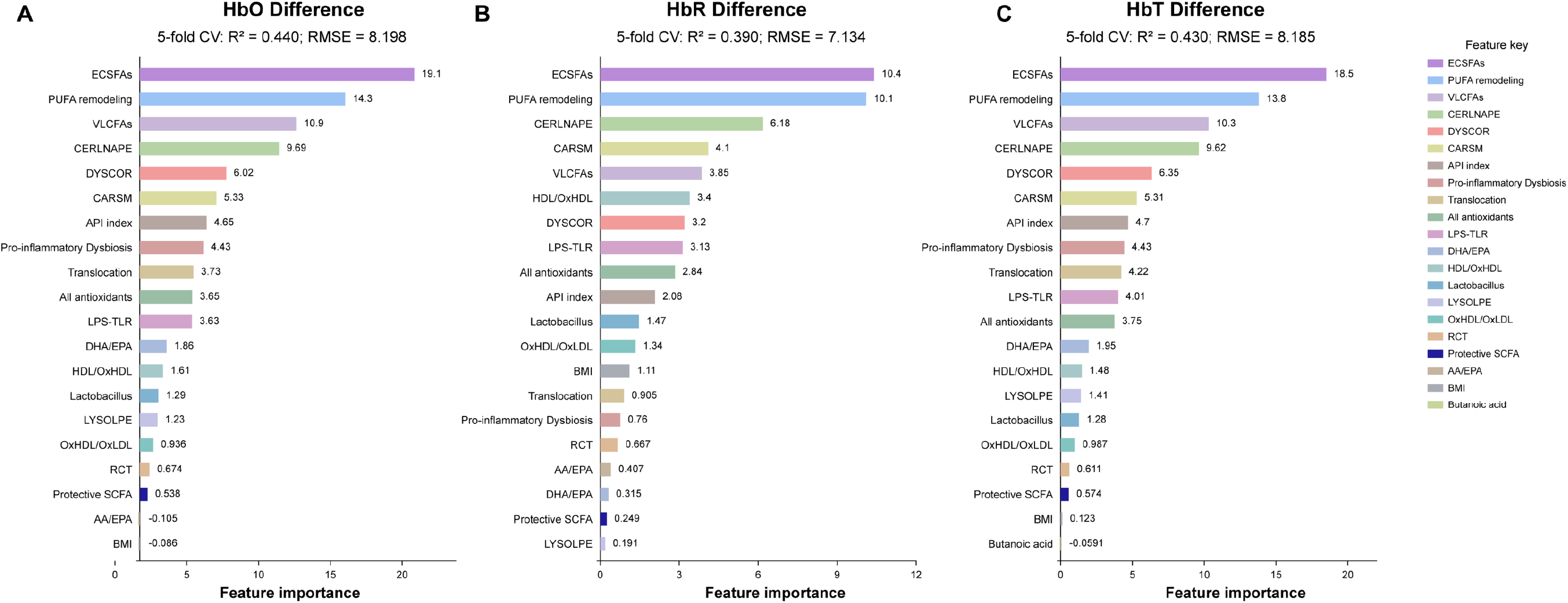
Peripheral NIMETOX predictors of fNIRS VFT Difference phenotypes. Random-forest regression models showing the relative importance of peripheral NIMETOX variables in explaining HbO (A), HbR (B), and HbT (C) VFT Difference composites. Model performance is expressed as cross-validated R² and RMSE. RF, random forest; RMSE, root mean square error. See ESF, Tables 1 and 2 for a detailed description of the NIMETOX biomarkers.

**Table 3** shows the results of multiple regression analyses. We found that 62.7% of the variance in HbO VFT Difference was explained by the model, with ECSFAs positively associated and age and the dysbiotic Coriobacteriia module (DYSCOR) inversely associated with HbO VFT Difference. In addition, 63.1% of the variance in HbR VFT Difference was explained by the model, with CERLNAPE, BMI, and age inversely associated with HbR VFT Difference. 64.1% of the variance in HbT VFT Difference was explained by the model, with ECSFAs positively associated and age, DYSCOR, and sex abuse inversely associated with HbT VFT Difference. For resting-state HbO Difference, 23.4% of the variance was explained by the model, with CARSM positively associated and age inversely associated with HbO Difference.

**Figure 6** shows that HbO, HbR and HbT Difference composites were significantly correlated with all key clinical domains and age, whereas there were no significant correlations with MetS or BMI. **Figure 7** shows the results of PLS and RF regression models. Panels 7B and 7C show that the regression of OSOD and the physiosomatic phenome, respectively, on the combined fNIRS and NIMETOX data yielded cross-validated R² values of 0.62 and 0.48. The most important explanatory variables in both models were ECSFAs, PUFA remodeling, All antioxidants, and DLPFC and FPA fNIRS measurements. The cross-validated R² values for RECUR (0.29) and Pure BDI (0.23) were more modest. The top variables contributing to the RECUR model were All antioxidants, gut microbiome Translocation, PUFA remodeling, Protective SCFAs, and CARSM. The fNIRS scores followed at a distance. The Pure BDI model was primarily predicted by DLPFC, FPA and S1 HbT Difference features, followed by PUFA remodeling.

**Figure 6.**
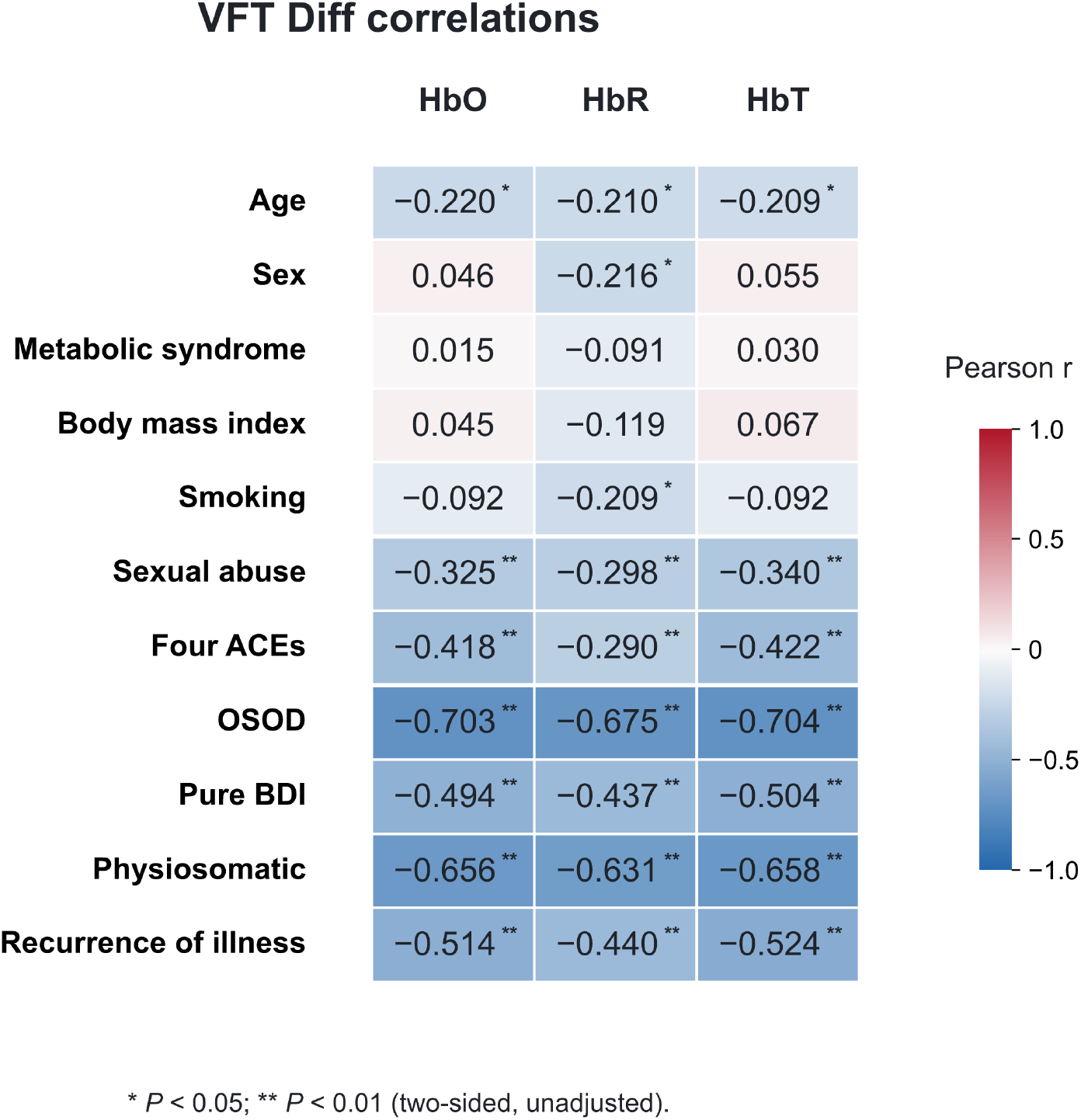
Associations of fNIRS VFT Difference phenotypes with clinical phenomes and demographic variables. Correlation matrix showing associations of HbO, HbR, and HbT VFT Difference composites with age, BMI, metabolic syndrome, and key clinical phenome scores, including OSOD, Pure BDI, the physiosomatic phenome, and recurrence of illness. OSOD, overall severity of depression; BMI, body mass index.

**Figure 7.**
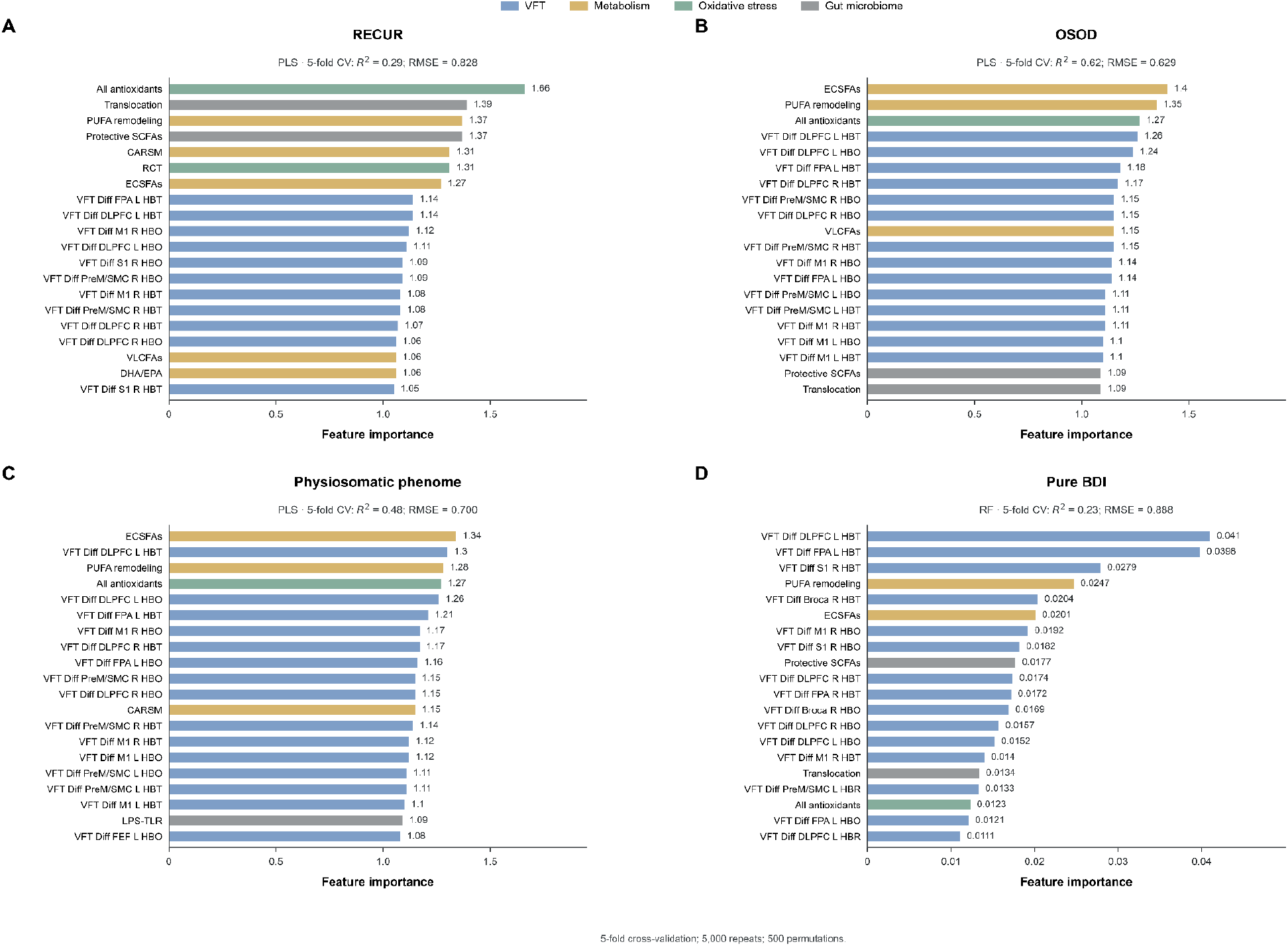
Integrated fNIRS–NIMETOX models explaining clinical phenome variation. Variable-importance profiles for models explaining (A) recurrence of illness (RECUR), (B) overall severity of depression (OSOD), (C) the physiosomatic phenome, and (D) Pure BDI using combined fNIRS Difference features and peripheral NIMETOX biomarkers. Model performance is expressed as cross-validated R² and RMSE. See ESF, Tables 1 and 2 for a detailed description of the NIMETOX biomarkers.

### 3.5 Variable-contribution profiles across HC and MDD patterns

**Figure 8** shows distinct variable-contribution profiles across HC and the three MDD patterns. HC showed positive contributions from VFT Difference and several metabolic and antioxidant variables, whereas all three MDD patterns showed negative VFT Difference contributions accompanied by distinct peripheral profiles. MDD pattern 1 (MDD1) was characterized by a prominent CERLNAPE contribution, MDD pattern 2 (MDD2) by negative ECSFAs and PUFA remodeling with positive gut-dysbiosis variables, and MDD pattern 3 (MDD3) by a prominent API and gut-microbiome contributions.

**Figure 8.**
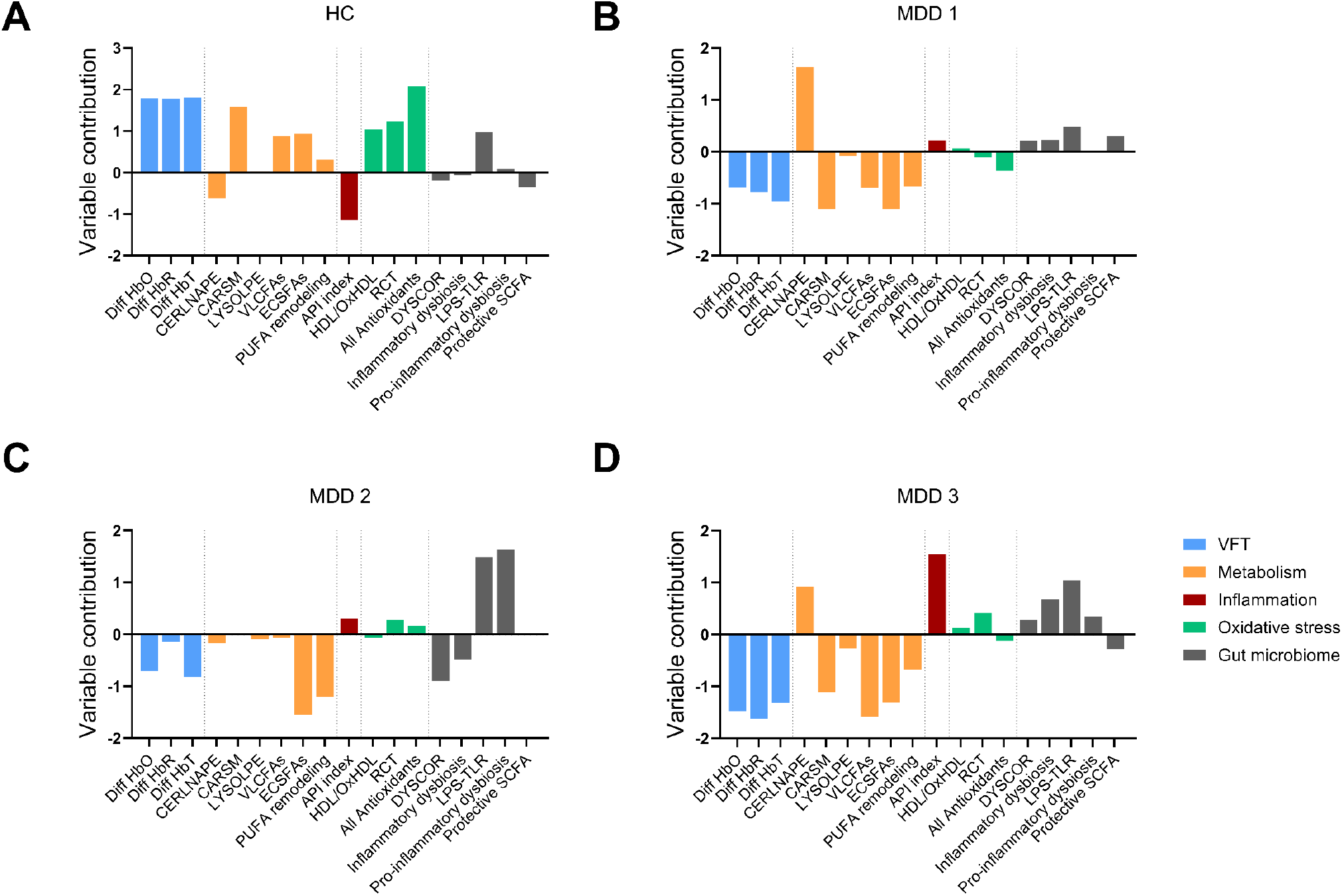
Variable-contribution profiles across one healthy control (HC) and three patients with major depressive disorder (MDD1–MDD3). Comparative variable-contribution profiles showing differences in fNIRS VFT Difference measures and peripheral NIMETOX features across HC and MDD1–MDD3. Positive and negative values indicate the direction of variable contributions within each profile.

## 4. Discussion

### 4.1 Hemodynamic differentiation of MDD and healthy controls using multivariate models

The first major finding of this study is that HbO, HbR, and HbT Difference and AUC were all significantly reduced during the VFT in patients with MDD, whereas resting-state abnormalities were comparatively limited, indicating that group differences in the measured fNIRS signals were more prominent under cognitive load. Previous fNIRS studies of MDD have repeatedly reported attenuated prefrontal and frontotemporal HbO responses during the VFT and other cognitive tasks^[10,56]^. Pu et al. found significantly smaller task-related hemodynamic changes in the prefrontal and temporal regions of patients with MDD than in healthy controls^[56]^. Kiriyama et al. reported significantly lower HbO changes across bilateral prefrontal and temporal regions during the VFT in MDD^[57]^. Dong et al. further demonstrated reduced prefrontal activation together with weaker overall and interhemispheric functional connectivity during the VFT^[58]^. In young adults with MDD, Kim et al. observed impaired oxygenation in prefrontal regions, including the frontopolar cortex, during VFT performance^[59]^. More recently, first-episode medication-naïve patients were shown to have significantly reduced activation in the frontal pole, inferior frontal gyrus, and dorsolateral prefrontal cortex during the VFT^[60]^. In a larger fNIRS dataset, Mao et al. also identified the frontopolar cortex, dorsolateral prefrontal cortex, and pars triangularis of Broca’s area among the most informative regions for differentiating MDD from healthy controls^[15]^. Collectively, these studies support a relatively consistent pattern of attenuated task-evoked prefrontal and frontotemporal hemodynamic responses in MDD, particularly during verbal fluency paradigms^[10]^.

The present findings are consistent with this general pattern but further show that MDD-related abnormalities encompass HbR, HbT dynamics, short-term temporal variability, and cumulative hemodynamic responses in addition to conventional HbO activation. “Difference” reflects the mean absolute change in hemodynamic signals between successive time points within a specified window; lower values indicate less point-to-point variation in the preprocessed signal^[61]^. AUC reflects cumulative hemodynamic output over a specified time window; its reduction indicates an attenuated overall hemodynamic response during the task^[13,35]^. Thus, the VFT abnormalities in MDD were not confined to a reduction at a single time point or in a single mean activation-amplitude measure but involved both reduced temporal hemodynamic dynamics and attenuated cumulative task-evoked recruitment. The concordant direction of abnormalities across HbO, HbR, and HbT further indicates that these changes were not restricted to a single oxygenation component but may involve oxygenated blood-flow supply, oxygen extraction, and local cerebral blood-volume regulation. The substantially stronger abnormalities during VFT than at rest suggest that cognitive challenge may expose latent limitations in cortical hemodynamic responsiveness that are less apparent under resting conditions.

ROI analyses further defined the spatial extent of the task-related abnormalities. Reduced VFT Difference was widely distributed across the bilateral DLPFC, FPA, FEF, PreM/SMC, M1, S1, and Broca’s area, encompassing coordinated cortical systems involved in cognitive control, attentional orienting, semantic retrieval, speech-motor preparation, and sensorimotor integration^[62–65]^. By contrast, VFT AUC abnormalities were concentrated primarily in the PreM/SMC, M1, S1, and Broca’s area, which are more directly involved in task execution and output^[64,65]^. The broader ROI alterations in Difference than in AUC suggest that short-term dynamic measures may be more sensitive to relatively subtle hemodynamic abnormalities, whereas deficient cumulative output is more prominent in the language and sensorimotor execution components of the VFT.

In a previous study, the anxiety spectrum was characterized by reduced Peak and increased Difference, whereas MDD in the present study showed concurrent reductions in Difference and AUC^[61]^. These contrasting patterns of temporal variability suggest distinct modes of neurovascular regulation in the two disorders. In anxiety states, despite a limited overall response amplitude, greater short-term temporal hemodynamic variability may indicate that the neurovascular system retains some dynamic regulatory capacity, representing an inefficient compensatory or hyperresponsive state^[61]^. By contrast, concurrent reductions in short-term variability and cumulative responses in MDD may reflect more widespread blunting of hemodynamic responses and reduced flexibility and compensatory reserve in neurovascular regulation. The transition from a “low-amplitude–high-Difference” pattern in the anxiety spectrum to a “low-response–low-Difference” pattern in MDD may therefore represent a shift from compensatory dynamic regulation toward a less responsive cortical hemodynamic state across different psychopathological conditions, potentially involving altered neurovascular and microcirculatory regulation^[17,31]^.

### 4.2 Peripheral NIMETOX profiles explain fNIRS hemodynamic phenotypes

The second major finding of this study is the significant association between fNIRS hemodynamic abnormalities and multiple peripheral NIMETOX features, with the strongest explanatory performance observed for VFT Difference. The most consistently important predictors primarily reflected lipid and fatty-acid metabolism, including ECSFAs, PUFA remodeling, CERLNAPE, VLCFAs, and CARSM, with additional contributions from the API index, antioxidant defenses, and gut-related modules. Collectively, these gut-NIMETOX-brain abnormalities indicate coordinated associations between cortical hemodynamic phenotypes and the lipid depletion (CARSM, ether lipids, Protective SCFAs, PUFA remodeling and VLCFAs) – lipotoxicity (increased ceramides-gangliosides) and other key NIMETOX pathways such as inflammation and lower antioxidant levels^[18,23,27]^. Notably, the VFT Difference models showed substantially greater explanatory performance than the VFT AUC and resting-state models, suggesting that peripheral NIMETOX dysregulation is more strongly associated with short-term cortical hemodynamic variability during cognitive engagement than with resting-state hemodynamics or cumulative task-evoked responses.

Lipid remodeling is particularly relevant because the imbalance between increased ceramides and GM3 gangliosides versus other lipids and fatty acids measured here may reflect alterations in membrane organization, cellular signaling, and NIMETOX pathways^[66]^. Ceramides impair endothelial protein kinase B (Akt)/endothelial nitric oxide synthase (eNOS) signaling, increase oxidative stress, reduce nitric oxide (NO) bioavailability, and compromise vascular responsiveness^[67,68]^. Increased GM3, a ceramide-derived glycosphingolipid, can disrupt receptor localization and signaling within membrane microdomains^[69]^ and inhibit vascular endothelial growth factor receptor 2 (VEGFR-2) signaling^[70]^. Thus, increased ceramide– GM3 metabolism may contribute to altered endothelial signaling, membrane function, and cortical hemodynamic responsiveness. The opposing relationships of ceramides/GM3 versus other lipids and fatty acids with fNIRS measures are compatible with increased partitioning toward lipotoxic sphingolipid pathways, potentially affecting mitochondrial, redox, and vascular functions^[27,71,72]^. Accordingly, reduced fNIRS responses were associated with a transition from structural and bioenergetic lipid and fatty-acid pools toward increased lipotoxicity^[23]^.

Excessive fatty-acid flux into sphingolipid pathways promotes ceramide accumulation and lipotoxicity, whereas circulating ECSFAs provide substrates for energy production and membrane lipid synthesis^[73,74]^. Thus, inverse associations between fNIRS measures and lipotoxic lipids may reflect detrimental fatty-acid partitioning, whereas positive associations with fatty acids, CARSM, and ether lipids may reflect preservation of membrane integrity and antioxidant defenses, mitochondrial fatty-acid transport and bioenergetics, acyl-coenzyme A (acyl-CoA) regulation, peroxisomal–membrane antioxidant functions, and substrates for pro-resolving pathways^[23,27]^. Low fatty-acid levels may also alter membrane composition, receptor organization, membrane microdomains, and eNOS-dependent vasodilation^[75]^. Saturated fatty acid (SFA) and PUFA status influences membrane fluidity, receptor function, neurotransmission, inflammation, and neuronal function^[76,77]^. Long-chain n-3 PUFA remodeling supports endothelial vasodilatory function^[78,79]^, while docosahexaenoic acid (DHA) supplementation can modify task-evoked HbO and HbT responses measured by fNIRS, directly linking fatty-acid status with cerebral hemodynamic responses during cognitive activation^[80]^.

Inflammation, reduced antioxidant defenses, and increased oxidative stress provide plausible mechanisms for the reduced VFT Difference observed in MDD. Inflammatory activation increases reactive oxygen species (ROS), while impaired antioxidant defenses favor oxidative and nitrosative stress, endothelial dysfunction, and disruption of the neurovascular unit. In particular, superoxide reduces NO bioavailability through its reaction with NO to form peroxynitrite, while oxidative stress can promote eNOS uncoupling, further increasing ROS production and impairing NO-dependent vascular regulation^[31]^. Because NO is a major mediator linking neuronal activation to local cerebral blood-flow responses, reduced NO bioavailability may impair neurovascular responses and the rapid microvascular adjustments required to meet changing metabolic demands during cognitive activation^[81,82]^. Consequently, the combined effects of inflammation, lowered antioxidant defenses, and oxidative stress may attenuate moment-to-moment vascular responsiveness during the VFT, resulting in smaller successive changes in HbO, HbR, and HbT, thereby lowering the Difference metric.

The gut-related findings suggest a complementary hypothesis involving the microbiota–gut– vascular–brain axis. Experimental disruption of the gut microbiota has been shown to impair cerebral arterial endothelial function and reduce eNOS activation, demonstrating that intestinal microbial composition can influence cerebral vascular physiology^[83]^. The DYSCOR module may be particularly relevant because *Eggerthella*, *Slackia*, and *Adlercreutzia* are specialized in transforming host- and diet-derived bioactive compounds, including polyphenols, isoflavones, and steroid-related metabolites. *Eggerthella lenta* has extensive capacity for biotransformation of endogenous and dietary small molecules and has also been linked to several chronic disease states^[84]^. *Slackia* and *Adlercreutzia* participate in isoflavone metabolism, including pathways generating equol and related metabolites with potential antioxidant and vascular actions^[85,86]^. Coriobacteriia-related changes may additionally interact with bile-acid metabolism, which regulates lipid metabolism, immune signaling, and host-microbiome communication^[87]^. Reduced *Lactobacillus* abundance may further indicate loss of protective microbiome functions involving intestinal-barrier maintenance, SCFA-related metabolism, and modulation of inflammatory and neuroendocrine signaling^[88]^.

### 4.3 fNIRS and NIMETOX biomarker profiles explain clinical phenome variation

The third major finding of this study is that integrated lipid-metabolic and fNIRS hemodynamic profiles showed good internal cross-validated differentiation between MDD and HC and explained substantial variation in key depressive phenotypes. PLS regression showed cross-validated explanatory performance for OSOD, the physiosomatic phenome, and RECUR, whereas RF regression yielded a more modest R² for Pure BDI. Across these models, lipid and fatty-acid metabolism, antioxidant-related variables, and fNIRS features from the frontopolar, dorsolateral prefrontal, premotor/sensorimotor, and Broca regions provided important explanatory information. These findings indicate that the major clinical dimensions of MDD are associated with an integrated NIMETOX–cortical hemodynamic profile rather than with isolated peripheral or cortical abnormalities.

The regional fNIRS findings are also functionally meaningful. FPA abnormalities may relate to higher-order cognitive integration and functional impairment, since reduced FPA VFT activation has previously been associated with impaired social functioning in MDD^[89]^. Moreover, impaired FPA oxygenation has been demonstrated in young adults with MDD and was particularly pronounced in patients with suicidality^[59]^. The contribution of PreM/SMC features may reflect alterations in cortical systems involved in action preparation, motor output, and the behavioral expression of cognitive effort, providing a plausible link with psychomotor and physiosomatic components of the phenome^[90,91]^. Broca-region involvement is particularly relevant to the VFT because this region participates in lexical retrieval, selection, and verbal production; reduced Broca-area HbO responses have been demonstrated in depressive states and associated with depression severity^[92]^. In a large MDD fNIRS study, Mao et al. likewise identified the FPA, DLPFC, and pars triangularis of Broca’s area among the most informative VFT regions^[15]^. Collectively, these findings suggest that OSOD and physiosomatic phenome variation is embedded within a distributed frontopolar–executive–language–motor hemodynamic phenotype coupled to peripheral phospholipid remodeling, lipotoxicity, fatty-acid dysregulation, and oxidative stress rather than being attributable to an isolated cortical region or metabolic pathway.

Importantly, the integrated models indicate that lipid-metabolic and cortical hemodynamic abnormalities provide partly complementary information on the clinical expression of MDD. Peripheral lipid abnormalities were associated with the fNIRS hemodynamic measures, supporting a potential pathway whereby disturbed lipid metabolism may influence the clinical phenome partly through alterations in cortical hemodynamic function. This interpretation is consistent with evidence that circulating lipid abnormalities are associated with cerebral blood-flow alterations in MDD^[93]^. However, the simultaneous retention of lipid-metabolic and fNIRS variables in the RF models suggests that the effects of lipid dysregulation and NIMETOX pathways on the MDD phenome are unlikely to be explained entirely by their associations with cortical hemodynamics. Thus, the current findings are compatible with a model in which lipid dysregulation may affect the MDD phenome through both hemodynamic and non-hemodynamic pathways, while altered cortical hemodynamic dynamics constitute one component of a broader NIMETOX-associated systems pathology.

### Limitations and strengths

Several limitations should be acknowledged. First, this was a cross-sectional study and therefore could not determine the temporal sequence or causal relationships among peripheral NIMETOX abnormalities, hemodynamic responses, and clinical phenotypes. Second, this study was designed as an explanatory systems-biology investigation rather than as a diagnostic classification study. Accordingly, the multivariate and machine-learning models were used primarily to identify and integrate cortical hemodynamic, peripheral NIMETOX, and clinical phenome relationships, rather than to establish diagnostic utility. Although internal cross-validation and bootstrap procedures supported the robustness of the identified models, their generalizability requires replication in independent cohorts, preferably across different clinical settings, ethnic and geographic populations, and fNIRS platforms. External validation will also be required before any diagnostic or clinical utility can be inferred. Third, although patients received different psychotropic medications, medication status did not materially affect the main findings, and no significant differences were observed between treated and untreated patients. This reduces the likelihood that medication exposure substantially accounted for the observed fNIRS and peripheral biomarker abnormalities. Nevertheless, medication dose, treatment duration, and cumulative exposure were not modeled and may exert more subtle effects among treated patients. These potential dose- and exposure-dependent effects should be examined in future studies specifically designed to address medication effects. In addition, dietary patterns, physical activity, sleep, and other metabolism-related factors in patients with MDD were not comprehensively assessed and may have influenced peripheral biomarker profiles and fNIRS hemodynamic measures. Finally, fNIRS primarily measures superficial cortical hemodynamics and cannot directly assess deep brain structures or the complete function of the neurovascular unit. The proposed cross-level pathway linking peripheral NIMETOX abnormalities, neurovascular function, cortical hemodynamics, and clinical phenotypes should therefore be regarded as a mechanistic hypothesis requiring further investigation.

A strength of this study was the use of 89-channel fNIRS to integrate resting-state and VFT conditions, HbO/HbR/HbT signals, and the two dynamic features Difference and AUC, together with multidimensional NIMETOX biomarkers and clinical phenomes. This approach enabled a systematic characterization of the neurovascular, peripheral biological, and clinically heterogeneous features of MDD.

## 5. Conclusions

In conclusion, our results acquired with an 89-channel fNIRS system revealed reductions in HbO, HbR, and HbT Difference and AUC during the VFT in MDD, whereas resting-state abnormalities were comparatively limited. These findings indicate lower point-to-point signal variation and smaller cumulative task-related measures across prefrontal, language, and sensorimotor regions. Peripheral NIMETOX profiles, particularly lipid and fatty-acid, oxidative-stress, SCFA, and gut-microbiota indices, explained substantial variation in fNIRS hemodynamic phenotypes, especially VFT Difference. Integrated fNIRS and peripheral biomarker profiles also explained variation in major clinical phenomes, particularly overall depression severity and physiosomatic symptoms. Thus, MDD is characterized by a multilevel systems-biological phenotype linking peripheral biological dysregulation, cortical hemodynamic abnormalities, and clinical heterogeneity. These findings support an explanatory systems-biology framework rather than established diagnostic utility. Independent longitudinal and multicenter studies are required to replicate these findings and establish their physiological and clinical significance.

## Abbreviations

AA/EPA: arachidonic acid/eicosapentaenoic acid ratio
acyl-CoA: acyl-coenzyme A
Akt: protein kinase B
AMP: amplitude threshold
API: acute-phase inflammatory
ApoA1: apolipoprotein A1
AUC: area under the concentration–time curve
BDI: Beck Depression Inventory
BMI: body mass index
CARSM: carnitine/sphingomyelin module
CERLNAPE: ceramide-, GM3 ganglioside-, N-acyl-phosphatidylethanolamine-, and phosphatidylethanolamine-enriched lipid component
CNS: central nervous system
mCRP: monomeric C-reactive protein
C-SSRS: Columbia-Suicide Severity Rating Scale
CV: coefficient of variation
DHA: docosahexaenoic acid
DLPFC: dorsolateral prefrontal cortex
DSM-5: Diagnostic and Statistical Manual of Mental Disorders Fifth Edition
DYSCOR: dysbiotic Coriobacteriia module
ECSFAs: even-chain saturated fatty acids
eNOS: endothelial nitric oxide synthase
ESF: Electronic Supplementary File
FDR: false discovery rate
FEF: frontal eye fields
FFS: FibroFatigue Scale
fNIRS: functional near-infrared spectroscopy
FPA: frontopolar area
GLM: general linear model
HAMA: Hamilton Anxiety Rating Scale
HAMD-21: 21-item Hamilton Depression Rating Scale
HbO: oxygenated hemoglobin
HbR: deoxygenated hemoglobin
HbT: total hemoglobin
HC: healthy controls
HDL: high-density lipoprotein
HDL-C: high-density lipoprotein cholesterol
LCAT: lecithin–cholesterol acyltransferase
LPS: lipopolysaccharide
M.I.N.I.: Mini-International Neuropsychiatric Interview
M1: primary motor cortex
MBLL: modified Beer–Lambert law
MDD: major depressive disorder
MetS: metabolic syndrome
MNI: Montreal Neurological Institute
MRI: magnetic resonance imaging
NIMETOX: neuroimmune: metabolic: and oxidative stress
NIRS-SPM: near-infrared spectroscopy statistical parametric mapping
NO: nitric oxide
OD: optical density
OPLS-DA: orthogonal partial least-squares discriminant analysis
OSOD: overall severity of depression
OxHDL: oxidized high-density lipoprotein
OxLDL: oxidized low-density lipoprotein
PCA: principal component analysis
PC1: principal component 1
PC2: principal component 2
PLS: partial least squares
PLS-DA: partial least-squares discriminant analysis
PON1: paraoxonase-1
PreM/SMC: premotor and supplementary motor cortex
PUFA(s): polyunsaturated fatty acid(s)
RCT: reverse cholesterol transport
RECUR: recurrence of illness
RF: random forest
RMSE: root mean square error
ROI/ROIs: region/regions of interest
ROS: reactive oxygen species
rRNA: ribosomal RNA
S1: primary somatosensory cortex
SCFA(s): short-chain fatty acid(s)
SFA: saturated fatty acid
SSS-8: Somatic Symptom Scale-8
STAI-State: State subscale of the State-Trait Anxiety Inventory
STDE: standard-deviation threshold
TAOC: total antioxidant capacity
TC: temporal cortex
TLR: Toll-like receptor
TMASK: masking window
TMOTION: motion-detection window
VEGFR-2: vascular endothelial growth factor receptor 2
VFT(s): verbal fluency task(s)
VIF: variance inflation factor
VIP: variable importance in projection
VLCFAs: very-long-chain fatty acids
WC: waist circumference
zBMI: standardized body mass index z score.

## Declarations

### Ethics approval and consent to participate

The study protocol was approved by the Ethics Committee of Sichuan Provincial People’s Hospital [Ethics (Research) 2024–203]. All participants or their legal representatives provided written informed consent before participation.

### Consent for publication

Not applicable.

### Availability of data and materials

De-identified data and analysis code may be made available from the corresponding author (MM) upon reasonable request and subject to institutional approval.

## Authors’ contributions

### Contributor Roles Taxonomy (CRediT)

**Yiping Luo:** Investigation, Resources, Data curation, Visualization, Writing – original draft, Writing – review & editing. **Xia Deng:** Investigation, Resources, Data curation, Writing – review & editing. **Mengqi Niu:** Investigation, Data curation, Writing – review & editing. **Andre F. Carvalho:** Writing – review & editing. **Hongzhou Wu:** Writing – review & editing. **Abbas F. Almulla:** Methodology, Formal analysis, Visualization, Writing – review & editing. **Xu Wang:** Investigation, Methodology, Writing – review & editing. **Jing Li:** Investigation, Resources, Writing – review & editing. **Yingqian Zhang:**Visualization, Writing – review & editing, Project administration, Funding acquisition. **Michael Maes:**

Conceptualization, Methodology, Validation, Formal analysis, Resources, Writing – review & editing, Supervision.

## Supporting information

esf

Graphical abstract

## Acknowledgements

None

