## Supplementary material for "Cortical hemodynamic dysregulation is linked to peripheral NIMETOX pathways in major depressive disorder: an 89-channel fNIRS systems-biology study": table

**Table 1. Demographic and clinical characteristics of patients with major depressive disorder (MDD) and healthy controls (HC)**

| Variable | HC (n = 40) | MDD (n = 86) | F/MWU/χ2 | df | p-value |
|---|---|---|---|---|---|
| Age (years) | 36.9 ± 13.7 | 34.3 ± 12.3 | F=1.11 | 1/124 | 0.295 |
| Gender (male/female) | 12 / 28 | 28 / 58 | $\chi^2$= 0.08 | 1 | 0.774 |
| Education (years) | 14.00 ± 4.32 | 13.51 ± 2.80 | F=0.58 | 1/124 | 0.447 |
| Body mass index (kg/m$^2$) | 23.65 ± 4.27 | 22.26 ± 3.26 | F=3.24 | 1/124 | 0.076 |
| Metabolic syndrome (yes/no) | 11/28 | 14/71 | $\chi^2$= 2.29 | 1 | 0.130 |
| Overall severity of depression (z scores) | -1.46 ± 0.26 | 0.59 ± 0.53 | MWU | - | <0.001 |
| Recurrence of illness (z score) | -1.06 ± 0.15 | 0.48 ± 0.93 | MWU | - | <0.001 |
| Pure BDI (z score) | -0.99 ± 0.46 | 0.44 ± 0.93 | MWU | - | <0.001 |
| Physiosomatic symptom domain (z score) | -1.30 ± 0.42 | 0.50 ± 0.66 | MWU | - | <0.001 |

**Note:** Continuous variables are presented as mean ± SD and categorical variables as counts. Categorical variables were compared using Pearson's chi-square test, and scale variables using analysis of variance or the Mann-Whitney U test (MWU). Pure BDI: the affective-cognitive symptoms of the Beck Depression Inventory.

**Table 2. Adjusted group differences in VFT and resting-state fNIRS indices**

| variable | Metric | Chromophore | HC adjusted mean ± SE | MDD adjusted mean ± SE | HC-MDD | F | df | *p*-value | FDR *q* |
|---|---|---|---|---|---|---|---|---|---|
| Rest | Difference | HbO | 0.386 ± 0.155 | -0.077 ± 0.105 | 0.463 | 6.469 | 1/118 | 0.012 | 0.021 |
| | | HbR | 0.003 ± 0.149 | -0.027 ±0.101 | 0.030 | 0.029 | 1/118 | 0.865 | 0.865 |
| | | HbT | 0.277 ± 0.156 | -0.039 ± 0.106 | 0.316 | 2.963 | 1/118 | 0.088 | 0.132 |
| | AUC | HbO | -0.012 ± 0.152 | 0.140 ± 0.103 | -0.152 | 0.732 | 1/118 | 0.394 | 0.473 |
| | | HbR | -0.089 ± 0.157 | 0.027 ±0.107 | -0.116 | 0.394 | 1/118 | 0.531 | 0.579 |
| | | HbT | 0.257 ± 0.151 | 0.041±0.103 | 0.216 | 1.481 | 1/118 | 0.226 | 0.301 |
| VFT | Difference | HbO | 1.074 ± 0.096 | -0.621 ± 0.073 | 1.694 | 210.651 | 1/100 | <0.001 | <0.001 |
| | | HbR | 1.100 ± 0.096 | -0.657 ±0.073 | 1.757 | 226.243 | 1/100 | <0.001 | <0.001 |
| | | HbT | 1.213 ± 0.097 | -0.579 ± 0.074 | 1.792 | 228.229 | 1/100 | <0.001 | <0.001 |
| | AUC | HbO | 0.546 ± 0.158 | -0.278 ± 0.120 | 0.824 | 18.328 | 1/100 | <0.001 | <0.001 |
| | | HbR | 0.545 ± 0.169 | -0.307 ± 0.128 | 0.852 | 17.053 | 1/100 | <0.001 | <0.001 |
| | | HbT | 0.615 ± 0.167 | -0.277 ± 0.127 | 0.892 | 19.183 | 1/100 | <0.001 | <0.001 |

**Note:** Values are adjusted mean ± SE from general linear models with diagnosis group and sex as fixed factors and MetS, age, and body mass index as covariates. FDR q values were calculated using the Benjamini–Hochberg procedure across the 12 MDD–HC comparisons. AUC, area under the curve; FDR, false discovery rate; VFT, verbal fluency task.

**Table 3. Results of multiple regression with fNIRS assessments as dependent variables and peripheral NIMETOX biomarkers as explanatory variables.**

| Dependent Variables | Explanatory Variables | Coefficients of input variables | | | Model statistics | | | |
|---|---|---|---|---|---|---|---|---|
| | | β | t | *p* | $R^2$ | F | df | *p* |
| #1 COMP HbO VFT Differences | **Model** | | | | 0.627 | 36.57 | 3/87 | < 0.001 |
| | Age | -0.308 | -4.67 | <0.001 | | | | |
| | ECSFAs | 0.310 | 3.93 | <0.001 | | | | |
| | DYSCOR | -0.179 | -2.60 | 0.011 | | | | |
| #2. COMP HbR VFT Differences | **Model** | | | | 0.631 | 29.42 | 3/86 | <0.001 |
| | CERLNAPE | -0.168 | -2.23 | 0.028 | | | | |
| | BMI | -0.185 | -2.71 | 0.008 | | | | |
| | Age | -0.175 | -2.50 | 0.014 | | | | |
| #3 COMP HbT VFT Differences | **Model** | | | | 0.641 | 30.65 | 4/86 | < 0.001 |
| | Age | -0.282 | -4.31 | <0.001 | | | | |
| | ECSFAs | 0.322 | 4.09 | <0.001 | | | | |
| | DYSCOR | -0.174 | -2.55 | 0.013 | | | | |
| | Sex abuse | -0.143 | -2.07 | 0.041 | | | | |
| #4 HbO Rest Differences | **Model** | | | | 0.234 | 13.60 | 2/89 | <0.001 |
| | Age | -0.370 | -3.97 | <0.001 | | | | |
| | CARSM | 0.282 | 3.03 | 0.003 | | | | |

**Note:** β, standardized regression coefficient; $R^2$, coefficient of determination; df, degrees of freedom. BMI, body mass index; CARSM, carnitine/sphingomyelin module; CERLNAPE, ceramide-centered lipid module; DYSCOR, dysbiotic Coriobacteriia-related module; ECSFAs, even-chain saturated fatty acids.
