## Supplementary material for "Cortical hemodynamic dysregulation is linked to peripheral NIMETOX pathways in major depressive disorder: an 89-channel fNIRS systems-biology study": esf

**ELECTRONIC SUPPLEMENTARY FILE (ESF)**

**ESF, Table 1. Analytical methods used to assess peripheral biomarkers in the present study**

| Biomarker domain / analyte | Analytical method and sample preparation | Equipment, software, and source |
|---|---|---|
| Untargeted metabolomics | Serum (50 µL) was precipitated with 300 µL acetonitrile:methanol (1:4, v/v) containing internal standards, vortexed for 3 min, centrifuged at 12,000 rpm for 10 min at 4°C, held at −20°C for 30 min, and recentrifuged. The supernatant was subjected to UPLC-HRMS in positive- and negative-ion modes; metabolites were annotated against KEGG, HMDB, and an in-house fragment library with a mass error <10 ppm. Metabolite data were reported as peak intensities and standardized z scores. | Vanquish UPLC coupled to Q Exactive HF-X Orbitrap HRMS; ACQUITY Premier HSS T3 column (2.1 × 100 mm, 1.8 µm); XCMS, CAMERA, and metaX [1] |
| Untargeted lipidomics | Lipids were extracted from 200 µL serum with 400 µL cold methyl tert-butyl ether and 80 µL cold methanol. After centrifugation, the supernatant was freeze-dried, reconstituted in dichloromethane:methanol (1:1), and analyzed by UPLC-HRMS. Features with excessive missingness were removed; remaining data underwent K-nearest-neighbor imputation, probabilistic quotient normalization, and centered log-ratio transformation. Among 1,095 lipid IDs, 157 met the differential-lipid criteria (ANOVA $p < 0.05$, fold change >1.20 or <0.83, and VIP >1). After removal of exogenous, drug-related, artifact-derived, technical-interference, and low-confidence signals, 111 lipids remained; LASSO, PLS-DA, and random forest analyses were then applied to define 43 featured lipids. Normalized lipid abundances were summarized as participant-level component scores. | ACQUITY UPLC CSH C18 column (2.1 × 100 mm, 1.7 µm) coupled to TripleTOF 6600 (AB SCIEX); XCMS, CAMERA, and metaX [2] |
| Fatty-acid profile | Targeted quantitative profiling of plasma fatty acids. Individual fatty-acid concentrations were used to derive chemical, structural, and ratio indices, including even-chain saturated fatty acids and the arachidonic acid/eicosapentaenoic acid ratio. Results were reported as concentrations and standardized composite or ratio scores. | Targeted plasma fatty-acid analytical protocol described in the source fatty-acid study [6] |
| 16S rRNA sequencing | Microbial DNA was extracted with the E.Z.N.A. Stool DNA Kit. The V4–V5 region of the bacterial 16S rRNA gene was amplified using primers 515F/907R and sequenced as paired-end 2 × 300-bp reads. Reads were filtered and assembled with Trimmomatic; OTUs were clustered at 97% similarity using UPARSE, chimeras were removed with UCHIME, and taxonomy was assigned using UCLUST against SILVA SSU138.1 at 80% confidence. Genus relative abundances were summarized as z-score composites. | Illumina amplicon sequencing; UPARSE, UCHIME, UCLUST, and SILVA SSU138.1 [3] |
| Monomeric C-reactive protein (mCRP) | Quantified by enzyme-linked immunosorbent assay (ELISA) and reported in ng/mL. Assay sensitivity was 0.63 ng/mL; intra-assay CV <10% and inter-assay CV <15%. | BioVendor ELISA kit, Brno, Czech Republic [5] |
| Albumin | Measured by immunoturbidimetry and reported in g/L. Analytical sensitivity was 0.03 g/L; intra-assay and inter-assay CVs were 1.96% and 0.67%, respectively. | DIAYS Diagnostic System reagent on an ADVIA 2400 analyzer [5] |
| Transferrin | Measured by the bromocresol green method and reported in g/L. Analytical sensitivity was 0.03 g/L; intra-assay and inter-assay CVs were 1.20% and 2.10%, respectively. | Beijing Strong Biotechnologies reagent on an ADVIA 2400 analyzer [5] |
| Short-chain fatty acids (SCFAs) | Targeted quantification after methanol extraction and 2-picolylamine derivatization with a deuterated internal standard. Derivatized extracts were separated by UPLC and quantified by tandem mass spectrometry in multiple-reaction-monitoring mode; calibration curves had $R^2$ >0.99. Results were expressed in µg/g feces or as rank-based standardized scores. | ACQUITY UPLC I-Class with BEH C18 column (2.1 × 100 mm, 1.7 µm) coupled to Xevo TQ-S triple-quadrupole MS; MassLynx 4.1 [4] |
| High-density lipoprotein cholesterol (HDL-C) | Measured using a direct selective-inhibition method and reported in mmol/L; intra-assay CV <4% and inter-assay CV <10%. | Beijing Strong Biotechnologies reagent on an ADVIA 2400 analyzer [5] |
| Oxidized HDL (OxHDL) | Quantified by ELISA and reported in ng/mL. Assay sensitivity was 0.938 ng/mL; intra-assay and inter-assay CVs were 5.12% and 5.22%, respectively. | FineTest EH4858 ELISA kit; Thermo Fisher SkyHigh microplate reader [5] |
| Oxidized LDL (OxLDL) | Quantified by ELISA and reported in pg/mL. Assay sensitivity was 37.5 pg/mL; intra-assay and inter-assay CVs were 6.62% and 7.04%, respectively. | Elabscience E-EL-H6021 ELISA kit; Thermo Fisher SkyHigh microplate reader [5] |
| TAOC | TAOC was quantified using a colorimetric assay kit. Assay sensitivity was 0.62 U/mL; intra-assay and inter-assay CVs were 4.8% and 5.6%, respectively. | Elabscience E-BC-K136-M kit (Wuhan, China); Thermo Fisher SkyHigh microplate reader [5] |
| PON1 (CMPAase) | Serum CMPAase activity was quantified by monitoring the hydrolysis of 4-(chloromethyl)phenyl acetate (CMPA; CAS No. 39720-27-9) at 280 nm in UV-transparent 96-well plates at 25 °C. Kinetic absorbance was recorded for 4 min (16 measurements at 15-s intervals), and activity was calculated in U/mL using a molar extinction coefficient of 1.30 mmol/L·cm−1. | CMPA substrate (Merck, USA); Thermo Fisher SkyHigh microplate reader [5] |

| Biomarker domain / analyte | Analytical method and sample preparation | Equipment, software, and source |
|---|---|---|
| LCAT index | Calculated as (1 − FC/TC) × 100 from free cholesterol (FC) and total cholesterol (TC). FC and TC were measured by cholesterol oxidase–phenol aminophenazone (CHOD-PAP) methods. | FC assay: mlbio, China; TC reagent: Beijing Strong Biotechnologies; ADVIA 2400 automated biochemical analyzer [8] |
| ApoA1 | Measured by immunoturbidimetry; intra-assay and inter-assay CVs were <3% and <10%, respectively. | Beijing Strong Biotechnologies reagent on an ADVIA 2400 automated biochemical analyzer [5] |

**Note:** Blood samples were collected under fasting conditions in the morning and stored at −80°C until analysis. CV, coefficient of variation; ELISA, enzyme-linked immunosorbent assay; HRMS, high-resolution mass spectrometry; LASSO, least absolute shrinkage and selection operator; PLS-DA, partial least-squares discriminant analysis; UPLC, ultra-performance liquid chromatography; VIP, variable importance in projection.

References [1–6,8] indicate the corresponding analytical-method source studies.

**ESF, Table 2.** Construction and harmonized nomenclature of biomarker composites used in the present study

| Biomarker domain | Label | Members/computation | Function / meaning | References |
|---|---|---|---|---|
| Lipidomics | CERLNAPE | 27 lipids, mainly ceramides, gangliosides, N-acyl lysophosphatidylethanolamines (LNAPE), and phosphatidylethanolamine (PE) species. | Ceramide-centered lipid module associated with lipotoxic signaling and membrane-lipid turnover; increased in the source MDD lipidomic study. | Zhang et al., 2026 [2] |
| Lipidomics | CARSM | Sphingomyelin 37:3;2O/7:0; CAR 22:0; CAR 10:0; 2-octenoylcarnitine; DL-acetylcarnitine. | Carnitine/sphingomyelin module related to mitochondrial fatty-acid handling and membrane homeostasis; lower scores indicate reduced bioenergetic support. | Zhang et al., 2026 [2] |
| Lipidomics | LYSOLPE | Lysophosphatidylcholine (LysoPC) (18:2/0:0); lysophosphatidylethanolamine (LysoPE) (P-18:0/0:0); lysophosphatidylethanolamine (LPE) 18:1; LPE O-20:1. | Lysophospholipid and ether-lysolipid module related to membrane turnover, repair, and antioxidant protection; depleted in the source MDD lipidomic study. | Zhang et al., 2026 [2] |
| Fatty acids | ECSFAs | C14:0, C16:0, C18:0, C20:0, C22:0, C24:0, C26:0. | Even-chain saturated fatty-acid pool supplying membrane phospholipids and ceramide precursors; depletion is interpreted in the context of structural-lipid remodeling. | Maes et al., 2026 [6] |
| Fatty acids | VLCFAs | C22:0, C22:2, C22:3, C22:4, C22:6n-3, C22:5n-6, C22:7, C23:0, C24:0, C24:1, C25:0, C26:0, C26:1. | Very-long-chain structural fatty acids involved in peroxisomal handling, and sphingolipid/myelin biology. | Maes et al., 2026 [6] |
| Fatty acids | PUFA remodeling | C16:2, C16:3, C16:4, C16:5, C17:2, C17:3, C18:4, C18:5, C19:2, C19:3, C19:4, C20:2, C20:6, C21:2, C21:3, C21:4, C21:5, C22:2, C22:3, C22:7. | Highly unsaturated fatty-acid pool indexing elongation–desaturation and membrane remodeling; relevant to oxidative susceptibility and depletion of membrane PUFA diversity. | Maes et al., 2026 [6] |
| Fatty acids | DHA/EPA | zC22:6n-3 - zC20:5n-3. | Relative abundance of DHA versus EPA within the long-chain n-3 fatty-acid pool. | Maes et al., 2026 [6] |

| Biomarker domain | Label | Members/computation | Function / meaning | References |
|---|---|---|---|---|
| Fatty acids | AA/EPA | zC20:4n-6 - zC20:5n-3 | Relative abundance of the n-6 precursor AA versus the n-3 precursor EPA; reflects the balance of fatty-acid substrates for lipid mediators. | Maes et al., 2026 [6] |
| Oxidized lipoproteins | OxHDL/OxLDL | zOxHDL − zOxLDL | Relative predominance of HDL versus LDL oxidation; a higher score indicates more OxHDL relative to OxLDL on the standardized scale. | Almulla et al., 2026 [5] |
| Oxidized lipoproteins | HDL/OxHDL | zHDL-C – zOxHDL | HDL availability relative to its oxidized fraction; a lower score indicates greater HDL oxidation susceptibility. | Almulla et al., 2026 [5] |
| Lipoprotein / redox | RCT | zHDL-C + zPON1 + zApoA1 + zLCAT | Reverse cholesterol transport and associated antioxidant functions; supports cholesterol efflux and return to the liver. | Almulla et al., 2023 [7] |
| Antioxidant defenses | All antioxidants | zAlbumin + zHDL-C + zApoA1 + zCMPAase + zTAOC | Combined circulating antioxidant defenses; lower values indicate reduced antioxidant capacity. | Almulla et al., 2026 [5] |
| Acute-phase response | API index | zmCRP − zalbumin − ztransferrin | Balance of a positive acute-phase reactant against two negative acute-phase proteins; higher values indicate a stronger acute-phase inflammatory response. | Almulla et al., 2026 [5] |
| Gut microbiome | DYSCOR | zEggerthella + zSlackia − zAdlercreutzia. | Actinobacterial functional polarization between opposing polyphenol-related metabolic roles. | Meng et al., 2026 [3] |
| Gut microbiome | Lactobacillus | zLactiplantibacillus + zLimosilactobacillus + zLoigolactobacillus. | Lactate-producing protective module linked to microbial cross-feeding, barrier support, and immune regulation. | Meng et al., 2026 [3] |
| Gut microbiome | Pro-inflammatory Dysbiosis | Fusobacterium + Porphyromonas + Clostridioides + Eggerthella + Bacteroides | Pro-inflammatory microbial signature associated with endotoxin exposure and proteolytic metabolism. | Meng et al., 2026 [3] |

| Biomarker domain | Label | Members/computation | Function / meaning | References |
|---|---|---|---|---|
| Gut microbiome | LPS-TLR | zBacteroides + zParabacteroides + zSutterella + zOdoribacter. | Gram-negative microbial load linked to LPS–TLR signaling and gut-derived immune activation. | Meng et al., 2026 [3] |
| Gut microbiome | Translocation | zFusobacterium + zPorphyromonas + zPhocea. | Oral pathobiont enrichment associated with mucosal inflammation, impaired barrier function, and oral–gut microbial translocation. | Meng et al., 2026 [3] |
| Fecal SCFAs | Butanoic acid | Butanoic acid (butyrate; C4:0) | A four-carbon microbial fermentation product associated with epithelial energy supply and barrier support. | Niu et al., 2026 [4] |
| Fecal SCFAs | Protective SCFAs | zAcetic acid + zPropionic acid + zButyric acid. | Combined measured fecal acetate, propionate, and butyrate profile representing protective saccharolytic fermentation. | Niu et al., 2026 [4] |

**Note:** Harmonized labels are used throughout the manuscript, tables, and figures; z denotes a standardized score.

**Abbreviations:** AA, arachidonic acid; API, acute-phase inflammatory index; DHA, docosahexaenoic acid; ECSFAs, even-chain saturated fatty acids; EPA, eicosapentaenoic acid; HDL, high-density lipoprotein; LPS, lipopolysaccharide; PUFA, polyunsaturated fatty acid; RCT, reverse cholesterol transport; SCFA, short-chain fatty acid; TLR, Toll-like receptor; VLCFAs, very-long-chain fatty acids.

**ESF, Table 3. Coverage of brain regions by the 89-channel fNIRS montage.**

| Region of interest (ROI) | Channels |
|---|---|
| Left DLPFC | CH05 CH06 CH12 CH19 CH26 CH27 CH32 CH18 |
| Right DLPFC | CH22 CH36 CH28 CH29 CH21 CH15 CH10 CH11 |
| Left FPA | CH01 CH02 CH07 CH13 |
| Right FPA | CH14 CH09 CH03 CH04 |
| Left FEF | CH33 CH41 CH42 |
| Right FEF | CH43 CH44 CH35 |
| Left TC | CH48 CH69 CH59 CH80 |
| Right TC | CH89 CH68 CH79 CH58 |
| Left PreM/SMC | CH38 CH39 CH40 CH50 CH51 CH52 CH62 CH63 CH73 CH49 |
| Right PreM/SMC | CH57 CH75 CH64 CH65 CH54 CH55 CH56 CH45 CH46 CH47 |
| Left M1 | CH84 CH61 CH72 CH83 |
| Right M1 | CH86 CH76 CH66 CH85 |
| Left Broca's area | CH31 CH16 CH17 CH25 |
| Right Broca's area | CH30 CH23 CH24 CH37 |
| Left S1 | CH81 CH82 CH70 CH71 CH60 |
| Right S1 | CH67 CH77 CH78 CH87 CH88 |
| Midline | CH08 CH20 CH34 CH53 CH74 |

**Note:** Eight bilateral cortical regions were represented by 16 lateralized ROIs. Five midline channels were excluded from lateralized ROI aggregation. DLPFC, dorsolateral prefrontal cortex; FPA, frontopolar area; FEF, frontal eye fields; TC, temporal cortex; PreM/SMC, premotor and supplementary motor cortex; M1, primary motor cortex; S1, primary somatosensory cortex.

**ESF, Table 4. Region-specific hemodynamic alterations in HbO, HbR, and HbT between MDD and healthy controls**

| Variables | Chromophore | ROI | Hemisphere | HC adjusted mean ± SE | MDD adjusted mean ± SE | F | p | FDR q |
|---|---|---|---|---|---|---|---|---|
| VFT_difference | HbO | DLPFC | L | 0.917 ± 0.115 | -0.578 ± 0.080 | 110.37 | <0.001 | <0.001 |
| | | | R | 0.865 ± 0.127 | -0.562 ± 0.089 | 82.34 | <0.001 | <0.001 |
| | | FPA | L | 0.731 ± 0.115 | -0.558 ± 0.080 | 81.92 | <0.001 | <0.001 |
| | | | R | 0.716 ± 0.152 | -0.465 ± 0.106 | 39.36 | <0.001 | <0.001 |
| | | FEF | L | 0.542 ± 0.135 | -0.481 ± 0.094 | 37.69 | <0.001 | <0.001 |
| | | | R | 0.487 ± 0.116 | -0.474 ± 0.081 | 45.03 | <0.001 | <0.001 |
| | | PreM/SMC | L | 0.636 ± 0.118 | -0.561 ± 0.083 | 66.79 | <0.001 | <0.001 |
| | | | R | 0.729 ± 0.130 | -0.529 ± 0.091 | 61.38 | <0.001 | <0.001 |
| | | M1 | L | 0.846 ± 0.150 | -0.561 ± 0.105 | 57.66 | <0.001 | <0.001 |
| | | | R | 0.903 ± 0.127 | -0.595 ± 0.089 | 90.81 | <0.001 | <0.001 |
| | | S1 | L | 0.719 ± 0.127 | -0.495 ± 0.089 | 59.21 | <0.001 | <0.001 |
| | | | R | 1.006 ± 0.153 | -0.522 ± 0.107 | 64.90 | <0.001 | <0.001 |
| | | Broca | L | 0.890 ± 0.161 | -0.444 ± 0.113 | 44.81 | <0.001 | <0.001 |
| | | | R | 0.731 ± 0.175 | -0.375 ± 0.122 | 26.06 | <0.001 | <0.001 |
| | HbR | DLPFC | L | 0.866 ± 0.121 | -0.653 ± 0.084 | 105.89 | <0.001 | <0.001 |
| | | | R | 0.679 ± 0.119 | -0.587 ± 0.083 | 75.15 | <0.001 | <0.001 |
| | | FPA | L | 0.543 ± 0.185 | -0.425 ± 0.129 | 18.23 | <0.001 | <0.001 |
| | | | R | 0.342 ± 0.179 | -0.433 ± 0.125 | 12.45 | 0.001 | 0.001 |
| | | FEF | L | 0.449 ± 0.119 | -0.526 ± 0.083 | 45.12 | <0.001 | <0.001 |
| | | | R | 0.405 ± 0.158 | -0.328 ± 0.110 | 14.33 | <0.001 | <0.001 |
| | | PreM/SMC | L | 0.493 ± 0.106 | -0.571 ± 0.073 | 67.83 | <0.001 | <0.001 |
| | | | R | 0.641 ± 0.134 | -0.566 ± 0.093 | 54.28 | <0.001 | <0.001 |
| | | M1 | L | 0.674 ± 0.155 | -0.563 ± 0.108 | 42.27 | <0.001 | <0.001 |
| | | | R | 0.618 ± 0.148 | -0.510 ± 0.103 | 38.67 | <0.001 | <0.001 |
| | | S1 | L | 0.539 ± 0.181 | -0.454 ± 0.126 | 19.95 | <0.001 | <0.001 |
| | | | R | 0.699 ± 0.180 | -0.442 ± 0.125 | 26.93 | <0.001 | <0.001 |
| | | Broca | L | 0.845 ± 0.167 | -0.462 ± 0.116 | 41.09 | <0.001 | <0.001 |
| | | | R | 0.510 ± 0.177 | -0.395 ± 0.123 | 17.47 | <0.001 | <0.001 |
| | HbT | DLPFC | L | 0.944 ± 0.111 | -0.610 ± 0.077 | 128.80 | <0.001 | <0.001 |
| | | | R | 0.889 ± 0.121 | -0.578 ± 0.085 | 95.73 | <0.001 | <0.001 |

| | | | | | | | | |
|---|---|---|---|---|---|---|---|---|
| | | FPA | L | 0.755 ± 0.114 | -0.583 ± 0.080 | 89.90 | <0.001 | <0.001 |
| | | | R | 0.726 ± 0.164 | -0.434 ± 0.115 | 32.48 | <0.001 | <0.001 |
| | | FEF | L | 0.530 ± 0.135 | -0.474 ± 0.094 | 36.25 | <0.001 | <0.001 |
| | | | R | 0.482 ± 0.112 | -0.498 ± 0.078 | 50.09 | <0.001 | <0.001 |
| | | PreM/SMC | L | 0.632 ± 0.117 | -0.548 ± 0.082 | 65.90 | <0.001 | <0.001 |
| | | | R | 0.707 ± 0.131 | -0.528 ± 0.092 | 58.09 | <0.001 | <0.001 |
| | | M1 | L | 0.837 ± 0.143 | -0.571 ± 0.100 | 63.63 | <0.001 | <0.001 |
| | | | R | 0.854 ± 0.129 | -0.560 ± 0.090 | 78.43 | <0.001 | <0.001 |
| | | S1 | L | 0.702 ± 0.127 | -0.484 ± 0.088 | 57.34 | <0.001 | <0.001 |
| | | | R | 0.945 ± 0.154 | -0.519 ± 0.108 | 58.62 | <0.001 | <0.001 |
| | | Broca | L | 0.891 ± 0.163 | -0.448 ± 0.114 | 44.23 | <0.001 | <0.001 |
| | | | R | 0.756 ± 0.174 | -0.382 ± 0.121 | 28.05 | <0.001 | <0.001 |
| VFT AUC | HbO | PreM/SMC | R | 0.313 ± 0.142 | -0.353 ± 0.099 | 14.36 | <0.001 | 0.004 |
| | | Broca | L | 0.486 ± 0.176 | -0.296 ± 0.123 | 12.85 | 0.001 | 0.004 |
| | HbR | PreM/SMC | L | 0.568 ± 0.176 | -0.282 ± 0.122 | 15.58 | <0.001 | 0.002 |
| | | M1 | L | 0.384 ± 0.166 | -0.370 ± 0.116 | 13.69 | <0.001 | 0.002 |
| | | | R | 0.316 ± 0.082 | -0.051 ± 0.057 | 13.35 | <0.001 | 0.002 |
| | HbT | Broca | R | 0.436 ± 0.192 | -0.243 ± 0.134 | 8.36 | 0.005 | 0.018 |
| | | PreM/SMC | R | 0.197 ± 0.135 | -0.344 ± 0.094 | 10.52 | 0.002 | 0.025 |
| | | S1 | L | 0.198 ± 0.118 | -0.207 ± 0.083 | 7.65 | 0.007 | 0.033 |
| | | Broca | L | 0.348 ± 0.164 | -0.241 ± 0.114 | 8.46 | 0.005 | 0.033 |

**Note:** Values are standardized adjusted means (z scores) ± SE from general linear models (df=1/76, except HbR: df=1/75). Only ROI-level results surviving Benjamini–Hochberg FDR correction ($q < 0.05$) within each chromophore × metric block are shown. All displayed results are from VFT models. FDR, false discovery rate; ROI, region of interest; VFT, verbal fluency task.

**ESF, Table 5. Group differences in peripheral biomarker composites between patients with major depressive disorder (MDD) and healthy controls (HC)**

| Biomarker composite (z scores) | HC adjusted mean (SE) n=39 | MDD adjusted mean (SE) n=86 | F | df | q (FDR) |
|---|---|---|---|---|---|
| API index | -1.516 (0.294) | 0.404 (0.198) | 28.91 | 1/ 119 | <0.001 |
| z OxHDL- z OxLDL | -0.574 (0.163) | 0.252 (0.110) | 17.36 | 1/119 | <0.001 |
| z HDL – z oxHDL | 0.592 (0.145) | -0.264 (0.098) | 23.67 | 1/119 | <0.001 |
| All antioxidants | 0.832 (0.143) | -0.330 (0.096) | 44.93 | 1/119 | <0.001 |
| RCT index | 0.600 (0.140) | -0.286 (0.095) | 27.02 | 1/119 | <0.001 |
| CERLNAPE | -0.930 (0.124) | 0.337 (0.083) | 70.99 | 1/120 | <0.001 |
| CARSM | 0.830 (0.128) | -0.282 (0.086) | 50.99 | 1/120 | <0.001 |
| LYSOLPE | 0.520 (0.149) | -0.315 (0.100) | 21.28 | 1/120 | <0.001 |
| ECSFAs | 1.326 (0.110) | -0.397 (0.073) | 168.50 | 1/120 | <0.001 |
| PUFAs | 1.286 (0.109) | -0.376 (0.073) | 158.16 | 1/120 | <0.001 |
| VLCFAs | 1.192 (0.118) | -0.302 (0.079) | 109.76 | 1/120 | <0.001 |
| z AA – z EPA | -0.064 (0.164) | 0.110 (0.110) | 0.767 | 1/120 | 0.383 |
| z DHA – z EPA | -0.624 (0.138) | 0.175 (0.093) | 22.71 | 1/120 | <0.001 |
| Butyric acid | 0.268 (0.156) | -0.141 (0.113) | 4.46 | 1/105 | 0.039 |
| Protective SCFAs | 1.145 (0.418) | -0.604 (0.302) | 11.35 | 1/105 | 0.0011 |
| Pro-inflammatory Dysbiosis | -0.613 (0.158) | 0.185 (0.114) | 16.58 | 1/105 | <0.001 |
| DYSCOR | -0.531 (0.158) | 0.169 (0.114) | 12.75 | 1/105 | <0.001 |
| Lactobacillus | 0.451 (0.164) | -0.236 (0.118) | 11.37 | 1/105 | 0.0011 |
| LPS-TLR | -0.764 (0.147) | 0.262 (0.107) | 31.32 | 1/105 | <0.001 |
| Translocation | -0.745 (0.148) | 0.243 (0.107) | 28.92 | 1/105 | <0.001 |

**Note:** Values are adjusted mean (SE) for age, sex and body mass index. See ESF, Tables 1-2 for explanation.
