## Supplementary figures and images for "Cortical hemodynamic dysregulation is linked to peripheral NIMETOX pathways in major depressive disorder: an 89-channel fNIRS systems-biology study"

### Graphical abstract

#### Graphical abstract

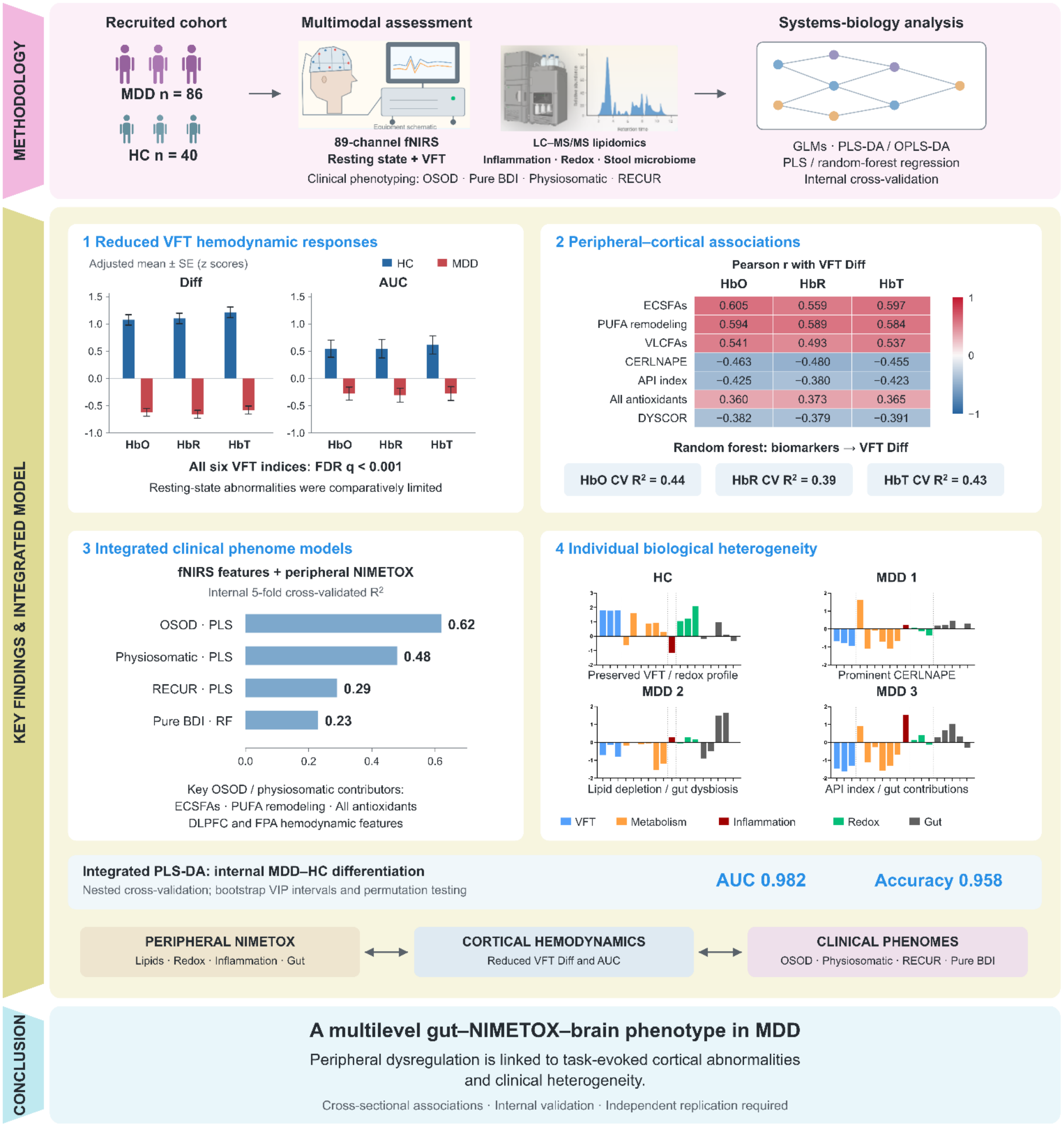
